# Use of U.S. Blood Donors for National Serosurveillance of SARS-CoV-2 Antibodies: Basis for an Expanded National Donor Serosurveillance Program

**DOI:** 10.1101/2021.05.01.21255576

**Authors:** Mars Stone, Clara Di Germanio, David J. Wright, Hasan Sulaeman, Honey Dave, Rebecca V. Fink, Edward P. Notari, Valerie Green, Donna Strauss, Debra Kessler, Mark Destree, Paula Saa, Phillip C. Williamson, Graham Simmons, Susan L. Stramer, Jean Opsomer, Jefferson M. Jones, Steven Kleinman, Michael P. Busch, for the NHLBI Recipient Epidemiology and Donor Evaluation Study-IV-Pediatric (REDS-IV-P)

**Affiliations:** Vitalant Research Institute (VRI), San Francisco, CA; Department of Laboratory Medicine, University of California, San Francisco, CA; Westat, Rockville, MD; American Red Cross (ARC), Gaithersburg, MD; Creative Testing Solutions (CTS), Tempe, AZ; New York Blood Center (NYBC), New York, NY; BloodWorks Northwest, Seattle, WA; Centers for Disease Control and Prevention COVID-19 Response Team, Atlanta, Georgia; University of British Columbia, Victoria, BC, Canada

**Keywords:** SARS-CoV-2, COVID-19, seroprevalence, COVID-19 Serological Testing

## Abstract

**Introduction:** The REDS-IV-P Epidemiology, Surveillance and Preparedness of the Novel SARS-CoV-2 Epidemic (RESPONSE) seroprevalence study conducted monthly cross-sectional testing for SARS-CoV-2 antibodies on blood donors in six U.S. metropolitan regions to estimate the extent of SARS-COV-2 infections over time.

**Study Design/Methods:** During March-August 2020, approximately ≥1,000 serum specimens were collected monthly from each region and tested for SARS-CoV-2 antibodies using a well-validated algorithm. Regional seroprevalence estimates were weighted based on demographic differences with the general population. Seroprevalence was compared with reported COVID-19 case rates over time.

**Results/Findings:** For all regions, seroprevalence was <1.0% in March 2020. New York experienced the biggest increase (peak seroprevalence, 15.8 % in May). All other regions experienced modest increases in seroprevalence(1-2% in May-June to 2-4% in July-August). Seroprevalence was higher in younger, non-Hispanic Black, and Hispanic donors. Temporal increases in donor seroprevalence correlated with reported case rates in each region. In August, 1.3-5.6 estimated cumulative infections (based on seroprevalence data) per COVID-19 case reported to CDC.

**Conclusion:** Increases in seroprevalence were found in all regions, with the largest increase in New York. Seroprevalence was higher in non-Hispanic Black and Hispanic blood donors than in non-Hispanic White blood donors. SARS-CoV-2 antibody testing of blood donor samples can be used to estimate the seroprevalence in the general population by region and demographic group. The methods derived from the RESPONSE seroprevalence study served as the basis for expanding SARS-CoV-2 seroprevalence surveillance to all 50 states and Puerto Rico.

**Summary:** SARS-CoV-2 serosurveillance data from blood donors in 6 US regions were used to estimate population weighted seroprevalence. Seroprevelance rates were higher in case rates. The study was expanded to a national donor serosurveillance program.

**Disclaimer:** The content is solely the responsibility of the authors and does not represent the policy of the National Institutes of Health or the Department of Health and Human Services. Any specific brand names included in this manuscript are for identification purposes only and are not intended to represent an endorsement by CDC. The findings and conclusions in this report are those of the authors and do not necessarily represent the official position of the Centers of Disease Control and Prevention.

## Introduction

Globally, as of January 2021, SARS-CoV-2 has caused nearly 100 million diagnosed COVID-19 cases, over two million deaths, and a substantial number of infections that are either asymptomatic or mildly symptomatic (1-3). With application of sensitive and specific serological assays and algorithms to representative populations, SARS-CoV-2 serosurveys are critical for estimating total infection rates, infection fatality rates, extent of herd immunity, and the effect of epidemic mitigation policies (4). Blood-donor-based serosurveillance is a powerful and cost-effective strategy that has provided valuable insights on infection prevalence and incidence for past emerging infectious threats including West Nile Virus, dengue, chikungunya and Zika (5-10). Choice of assays for serosurveillance should be determined by intended purpose (11, 12) and assay performance which can be influenced by antigen and immunoglobulin targets, and assay configuration (13).

In response to the emergence of COVID-19 in the United States in early 2020, the National Heart, Lung and Blood Institute Recipient Epidemiology and Donor Evaluation Study (REDS-IV-P) program developed and implemented molecular and serologic surveillance for SARS-CoV-2 in six metropolitan regions, called the **R**EDS-IV-P **E**pidemiology, **S**urveillance and **P**reparedness of the **N**ovel **S**ARS-CoV-2 **E**pidemic (RESPONSE) study. RESPONSE project aims included conducting testing for SARS-CoV-2 antibodies to estimate seroprevalence, to evaluate trends in seroprevalence, and to compare the observed seroprevalence with reported case data.

## METHODS

### Study Sites and Donation Sampling

The RESPONSE study tested for SARS-CoV-2 antibodies in three early-outbreak regions starting in March 2020 (Seattle, New York, and San Francisco), and three initially low-prevalence regions in April 2020 (Boston, Los Angeles, and Minneapolis)(see Table 1 for donor characteristics and Figure 1 for testing algorithm). About 1000 serum specimens were randomly selected monthly from allogeneic blood donors from March/April through August 2020. In July and August, monthly sampling increased to 2000-4000 per region as the study transitioned into the expanded Multistate Assessment of SARS-CoV-2 Seroprevalence in Blood Donors (MASS-BD) Study (14). Beginning in June 2020, the blood collection organizations associated with four regions (San Francisco, Los Angeles, Minneapolis, and Boston) began screening all blood donors for SARS-CoV-2 antibodies.(15) In July and August in these regions, antibody data were extracted from donation records, whereas for Seattle and New York, study-initiated testing continued. For all months, donations made specifically to provide COVID-19 convalescent plasma (CCP) were excluded. The study was reviewed by the UCSF Committee for Human Research and determined to meet the definition of research as defined in 46.102(l) but did not involve human subjects based on anonymization of data and routine consent for blood donation testing that includes use of residual samples for research purposes (as defined in 46.103(e)(1) consistent with applicable federal law and CDC policy (45 C.F.R. part 46; 21 C.F.R. part 56; 42 U.S.C. §241(d), 5 U.S.C. §552a, 44 U.S.C. §3501)). We used the STROBE cross sectional checklist when writing our report. (16)

**Table 1.** Demographic characteristics of donors that provided specimens included in study and of all donors who donated in each region, six U.S. metropolitan regions, March–August 2020. Rh = rhesus factor.

|  |  | New York |  | San Francisco |  | Seattle |  | Boston |  | Los Angeles |  | Minneapolis |  | All Regions |  |
| --- | --- | --- | --- | --- | --- | --- | --- | --- | --- | --- | --- | --- | --- | --- | --- |
|  |  | All Donations | Sampled Donations | All Donations | Sampled Donations | All Donations | Sampled Donations | All Donations | Sampled Donations | All Donations | Sampled Donations | All Donations | Sampled Donations | All Donations | Sampled Donations |
| All |  | 131622 | 9132 | 28758 | 7986 | 76209 | 8019 | 47437 | 6999 | 114692 | 11000 | 98010 | 7000 | 496728 | 50156 |
|  | Gender |  |  |  |  |  |  |  |  |  |  |  |  |  |  |
|  | Female | 46.5 | 47.5 | 51.6 | 50.8 | 56.7 | 55.7 | 53.6 | 53.8 | 54.7 | 54.1 | 57.5 | 57.8 | 53.1 | 53.1 |
|  | Male | 53.5 | 52.5 | 48.4 | 49.2 | 43.3 | 44.3 | 46.4 | 46.2 | 45.3 | 45.9 | 42.5 | 42.2 | 46.9 | 46.9 |
| Age |  |  |  |  |  |  |  |  |  |  |  |  |  |  |  |
|  | 16-29 | 18.0 | 20.5 | 12.5 | 11.8 | 14.4 | 13.1 | 13.6 | 14.4 | 16.3 | 16.4 | 10.8 | 9.8 | 14.9 | 14.7 |
|  | 30-49 | 30.2 | 31.2 | 30.8 | 32.0 | 34.2 | 32.6 | 28.5 | 29.1 | 35.5 | 36.3 | 28.2 | 29.0 | 31.5 | 32.1 |
|  | 50-64 | 38.8 | 34.5 | 36.9 | 36.1 | 32.2 | 32.6 | 40.3 | 39.2 | 34.5 | 34.3 | 36.1 | 36.7 | 36.3 | 35.4 |
|  | 65+ | 13.0 | 13.8 | 19.8 | 20.2 | 19.2 | 21.7 | 17.6 | 17.2 | 13.7 | 13.1 | 24.9 | 24.5 | 17.3 | 17.9 |
| Race/<br>Ethnicity |  |  |  |  |  |  |  |  |  |  |  |  |  |  |  |
|  | White | 78.5 | 77.3 | 71.3 | 71.3 | 81.2 | 83.8 | 92.6 | 92.7 | 62.4 | 60.5 | 97.0 | 97.0 | 79.8 | 78.6 |
|  | Black | 3.6 | 3.8 | 1.4 | 1.4 | 0.9 | 0.7 | 1.0 | 1.0 | 2.1 | 2.2 | 0.3 | 0.3 | 1.8 | 1.7 |
|  | Hispanic | 8.9 | 8.0 | 8.2 | 8.4 | 2.4 | 2.3 | 2.0 | 2.2 | 19.9 | 20.8 | 0.8 | 0.7 | 8.1 | 8.1 |
|  | Other | 8.9 | 10.9 | 19.2 | 18.8 | 15.6 | 13.2 | 4.4 | 4.1 | 15.7 | 16.4 | 1.9 | 2.0 | 10.3 | 11.6 |
| Blood<br>group |  |  |  |  |  |  |  |  |  |  |  |  |  |  |  |
|  | O | 50.9 | 49.4 | 48.6 | 48.2 | 50.4 | 52.0 | 51.2 | 51.6 | 50.7 | 50.6 | 47.5 | 47.3 | 50.0 | 49.9 |
|  | A | 32.6 | 33.5 | 33.7 | 34.4 | 35.3 | 34.9 | 33.2 | 33.1 | 32.8 | 32.4 | 37.3 | 37.0 | 34.1 | 34.1 |
|  | B | 12.3 | 12.5 | 12.2 | 12.4 | 10.3 | 9.9 | 11.2 | 10.7 | 11.9 | 12.3 | 10.6 | 11.0 | 11.4 | 11.6 |
|  | AB | 4.2 | 4.5 | 5.5 | 5.0 | 4.1 | 3.1 | 4.4 | 4.6 | 4.6 | 4.7 | 4.6 | 4.6 | 4.4 | 4.4 |
| Rh type |  |  |  |  |  |  |  |  |  |  |  |  |  |  |  |
|  | Rh positive | 83.3 | 84.2 | 83.0 | 83.5 | 79.9 | 79.2 | 80.7 | 80.5 | 84.7 | 84.5 | 78.5 | 77.9 | 81.9 | 81.9 |
|  | Rh negative | 16.7 | 15.8 | 17.0 | 16.5 | 20.1 | 20.7 | 19.3 | 19.4 | 15.3 | 15.5 | 21.4 | 22.1 | 18.1 | 18.0 |
| Donor<br>Status |  |  |  |  |  |  |  |  |  |  |  |  |  |  |  |
|  | First time | 14.6 | 14.8 | 28.4 | 18.0 | 21.1 | 16.2 | 22.8 | 21.8 | 30.9 | 27.3 | 18.0 | 16.2 | 21.6 | 19.5 |
|  | Repeat | 85.4 | 85.2 | 71.6 | 82.0 | 78.9 | 83.8 | 77.2 | 78.2 | 69.1 | 72.7 | 82.0 | 83.8 | 78.4 | 80.5 |

**Figure 1.**
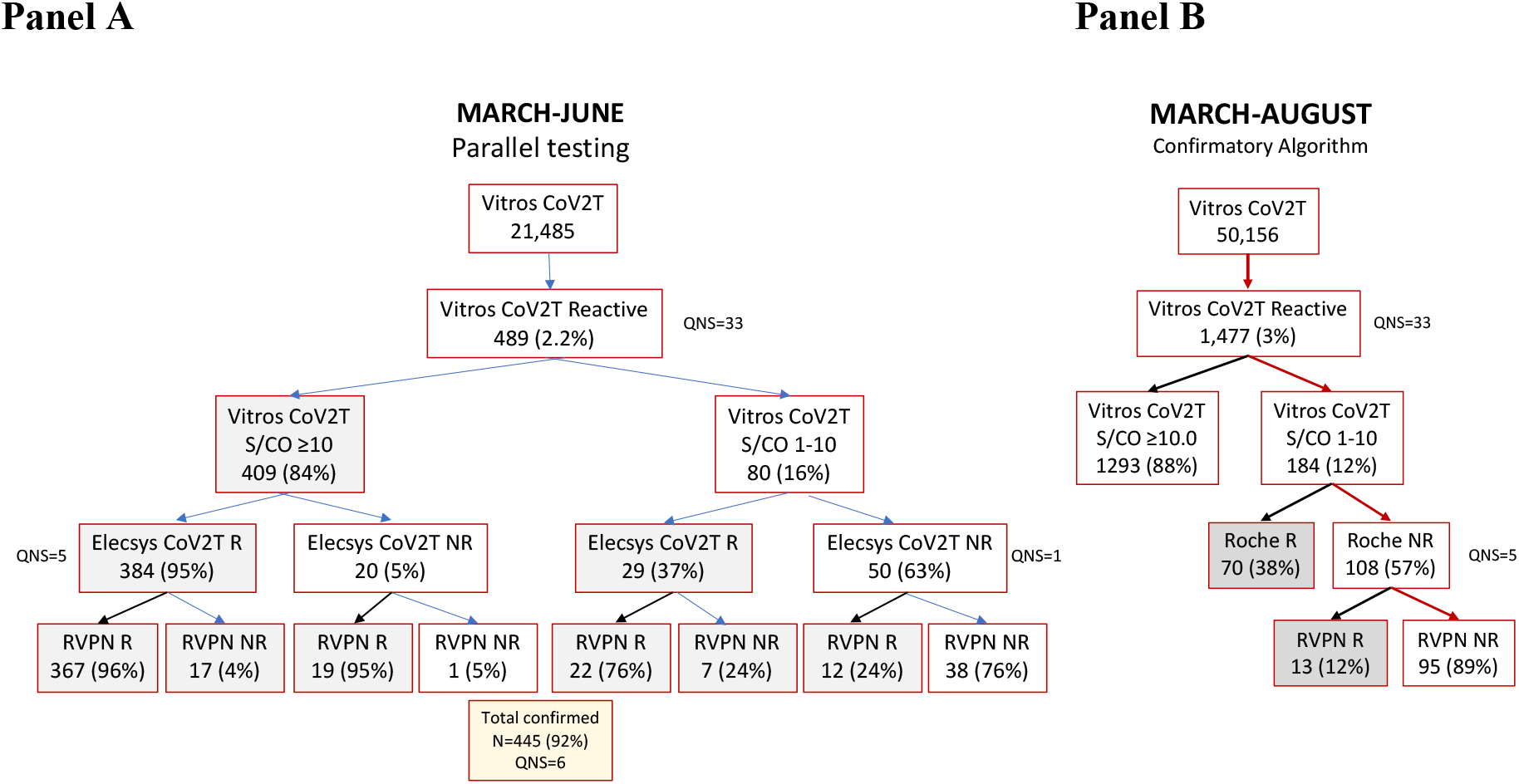
SARS-CoV-2 serology testing results flow chart, six U.S. metropolitan regions. Panel A: Parallel testing using Elecsys CoV2T and RVNPT assay on Ortho Vitros S1 Total Ig CoV2T reactive samples collected during March–June 2020. Panel B: Results from March through August 2020 combining the initial and revised supplementary testing algorithms. Vitros COV2T: Ortho VITROS Immunodiagnostic Products Anti-SARS-CoV-2 Total; QNS: quantity not sufficient; Elecsys CoV2T: Roche Elecsys Nucleocapsid Anti-SARS-CoV-2 Total Immunoglobulin; RVPN: pseudovirus reporter virus particle neutralization; R: reactive; NR: nonreative.

### Screening and supplemental serology assays and establishing a testing algorithm

Initially, the serology screening and supplemental testing algorithm consisted of screening all samples with the Ortho VITROS Immunodiagnostic Products Anti-SARS-CoV-2 Total test (Vitros CoV2T). Reactive samples were confirmed by parallel testing by both a nucleocapsid (NC)-based Total Ig assay (Roche Elecsys NC Anti-SARS-CoV-2 Total Ig) (Elecsys CoV2T) and a pseudovirus reporter virus particle neutralization test (RVPN) (Appendix A). Screened-positive specimens were considered confirmed if reactive by either Elecsys CoV2T or RVPN. The Vitros CoV2T and Elecsys CoV2T assays were selected based on their double antigen-sandwich design, which enables durable detection of total Ig and employed as an orthogonal algorithm to detect antibodies to different SARS-CoV-2 antigens (S1 and NC, respectively). FDA EUA Instructions for Use (IFU) (17) and other reports have demonstrated excellent sensitivity of both assays during acute infection and stability of antibody reactivity on serial samples collected >120 days from COVID-19 symptom onset. (18-20)

### Statistical methods to extrapolate donor seroprevalence to the general population

The geographic distribution and demographic composition of sampled donors varied monthly. To ensure sample populations represented a consistent geographic area over the course of the study, donations were restricted to ZIP codes in which at least 80% of donors resided, referred to in this study as the Donor Catchment Regions (DCRs). Donations from donors that resided outside of the DCR were excluded. Monthly sample donor demographics were compared with monthly total donation demographics at each blood center via Chi-square statistics (without accounting for a multiple comparison adjustment) to ensure that sampled donations were representative of general donor populations.

To estimate the monthly seroprevalence in the general population based on blood donor seroprevalence, monthly estimation weights were created that accounted for demographic difference between the blood donor sample and general population. The 2018 American Community Survey (ACS) estimates (21) for the age, gender, and race/ethnicity composition for the DCRs were used to standardize DCR sample totals by raking. In addition to these estimation weights, monthly sets of 50 pseudo-replicate weights were created to compute weighted seroprevalence standard errors. Because seroprevalence in the U.S. population is known to vary by location and time, a stratified (by blood center and month) logistic regression model was developed to assess the association between seropositivity and demographic characteristics.

Blood donation DCRs were defined by ZIP codes, but case reporting by state and local health departments to CDC is reported by county. Therefore, to compare the number of cumulative infections estimated from seroprevalence with the number of cumulative cases reported to CDC by each region, we created county-based DCRs. The number of total cumulative infections in a DCR was estimated by multiplying the weighted seroprevalence by the total population in the DCR. (See Supplemental Figure 1 and Appendix B for detailed statistical methods). For each county-based DCR, the number of cumulative infections based on seroprevalence was divided by the number of reported cases.

## RESULTS

### Validation of supplemental testing algorithm

During March-June 2020, a total of 21,485 donations were screened with Vitros CoV2T, of which 489 reactive specimens were tested in parallel by the Elecsys CoV2T and RVPN **(Figure 1a)**. Specimens were stratified based on Vitros CoV2T signal to cutoff (S/CO) ratios: specimens with S/CO 1-10 and specimens with S/CO ≥10. Parallel testing of all screened reactive specimens demonstrated that among the 404 specimens with Vitros CoV2T S/CO ≥10.0 and available Elecsys CoV2T results, 384 were Elecsys CoV2T reactive and 19 reactive by RVPN, thus >99% of specimens with Vitros CoV2T S/CO≥10 were confirmed reactive by either Elecsys CoV2T or RVPN. In contrast, of 79 screened reactive specimens with Vitros CoV2T S/CO 1−10 and available Elecsys CoV2T results, 29 were Elecsys CoV2T reactive and 12 were RVPN reactive, only 51% of specimens with S/CO 1-10 were confirmed reactive **(Figure 1a)**. Thus, beginning in July we modified the supplemental testing algorithm to be more cost-effective while maintaining high sensitivity and specificity for July and August **(Figure 1b)** so that specimens were considered “confirmed antibody positive” if: i) they had a S/CO ≥10 on Vitros CoV2T screening assay (i.e., no supplemental testing was performed); or ii) if the Vitros CoV2T S/CO was 1−10 and reactive on either the Elecsys CoV2T or RVPN assay. Details and results of application of this testing algorithm for the entire study interval (March-August) are presented in Appendix C.

### Seroprevalence estimates over time, with and without supplemental testing and population weighting

In total, 499,476 non-COVID-19 convalescent plasma donations were collected in all participating regions during the study period, of which 50,156 (10%) were included in the study. Monthly distributions of Vitros CoV2T reactivity, supplemental testing status, and number of tested specimens are shown in **Figure 2, Panel A**, and seroprevalence by month and site are presented in Supplemental Table 2. Low rates of unweighted confirmed seroreactivity (<1%) were observed for all regions at the beginning of the testing period in March 2020, with variable increases over the 5-6-month serosurveillance period. The greatest increase in seroprevalence was seen in New York (0.7% to 15.7%) followed by Los Angeles (0.8% to 4.5%) and Boston (0.9% to 4.2%). Mean Vitros CoV2T signal intensity increased from a S/CO of 37.8 (range: 1.1-182.4) in March to 308.9 (range: 1.0-1380.0) in August, demonstrating that both proportions of confirmed seropositive donations and mean signal intensities increased over time in each region.

**Table 2.**
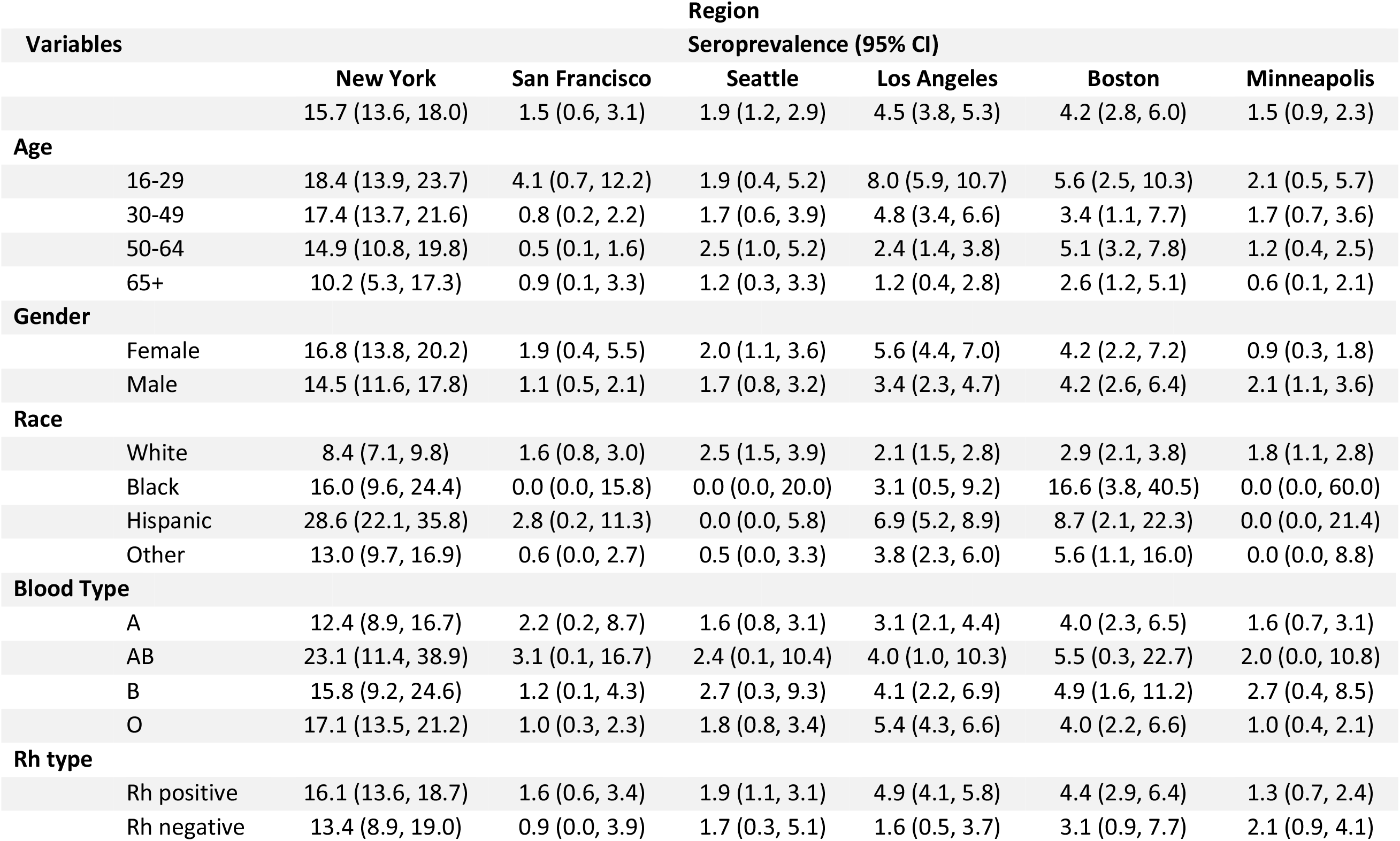
Weighted confirmed seroprevalence by demographic characteristics, six U.S. metropolitan regions, August 2020. 95% CI = 95% confidence interval; DCR = donor catchment region; Rh +ve = rhesus factor positive; Rh-ve = rhesus factor negative

| Variables |  | Region |  |  |  |  |  |
| --- | --- | --- | --- | --- | --- | --- | --- |
|  |  | Seroprevalence (95% CI) |  |  |  |  |  |
|  |  | New York | San Francisco | Seattle | Los Angeles | Boston | Minneapolis |
|  |  | 15.7 (13.6, 18.0) | 1.5 (0.6, 3.1) | 1.9 (1.2, 2.9) | 4.5 (3.8, 5.3) | 4.2 (2.8, 6.0) | 1.5 (0.9, 2.3) |
| Age |  |  |  |  |  |  |  |
|  | 16-29 | 18.4 (13.9, 23.7) | 4.1 (0.7, 12.2) | 1.9 (0.4, 5.2) | 8.0 (5.9, 10.7) | 5.6 (2.5, 10.3) | 2.1 (0.5, 5.7) |
|  | 30-49 | 17.4 (13.7, 21.6) | 0.8 (0.2, 2.2) | 1.7 (0.6, 3.9) | 4.8 (3.4, 6.6) | 3.4 (1.1, 7.7) | 1.7 (0.7, 3.6) |
|  | 50-64 | 14.9 (10.8, 19.8) | 0.5 (0.1, 1.6) | 2.5 (1.0, 5.2) | 2.4 (1.4, 3.8) | 5.1 (3.2, 7.8) | 1.2 (0.4, 2.5) |
|  | 65+ | 10.2 (5.3, 17.3) | 0.9 (0.1, 3.3) | 1.2 (0.3, 3.3) | 1.2 (0.4, 2.8) | 2.6 (1.2, 5.1) | 0.6 (0.1, 2.1) |
| Gender |  |  |  |  |  |  |  |
|  | Female | 16.8 (13.8, 20.2) | 1.9 (0.4, 5.5) | 2.0 (1.1, 3.6) | 5.6 (4.4, 7.0) | 4.2 (2.2, 7.2) | 0.9 (0.3, 1.8) |
|  | Male | 14.5 (11.6, 17.8) | 1.1 (0.5, 2.1) | 1.7 (0.8, 3.2) | 3.4 (2.3, 4.7) | 4.2 (2.6, 6.4) | 2.1 (1.1, 3.6) |
| Race |  |  |  |  |  |  |  |
|  | White | 8.4 (7.1, 9.8) | 1.6 (0.8, 3.0) | 2.5 (1.5, 3.9) | 2.1 (1.5, 2.8) | 2.9 (2.1, 3.8) | 1.8 (1.1, 2.8) |
|  | Black | 16.0 (9.6, 24.4) | 0.0 (0.0, 15.8) | 0.0 (0.0, 20.0) | 3.1 (0.5, 9.2) | 16.6 (3.8, 40.5) | 0.0 (0.0, 60.0) |
|  | Hispanic | 28.6 (22.1, 35.8) | 2.8 (0.2, 11.3) | 0.0 (0.0, 5.8) | 6.9 (5.2, 8.9) | 8.7 (2.1, 22.3) | 0.0 (0.0, 21.4) |
|  | Other | 13.0 (9.7, 16.9) | 0.6 (0.0, 2.7) | 0.5 (0.0, 3.3) | 3.8 (2.3, 6.0) | 5.6 (1.1, 16.0) | 0.0 (0.0, 8.8) |
| Blood Type |  |  |  |  |  |  |  |
|  | A | 12.4 (8.9, 16.7) | 2.2 (0.2, 8.7) | 1.6 (0.8, 3.1) | 3.1 (2.1, 4.4) | 4.0 (2.3, 6.5) | 1.6 (0.7, 3.1) |
|  | AB | 23.1 (11.4, 38.9) | 3.1 (0.1, 16.7) | 2.4 (0.1, 10.4) | 4.0 (1.0, 10.3) | 5.5 (0.3, 22.7) | 2.0 (0.0, 10.8) |
|  | B | 15.8 (9.2, 24.6) | 1.2 (0.1, 4.3) | 2.7 (0.3, 9.3) | 4.1 (2.2, 6.9) | 4.9 (1.6, 11.2) | 2.7 (0.4, 8.5) |
|  | O | 17.1 (13.5, 21.2) | 1.0 (0.3, 2.3) | 1.8 (0.8, 3.4) | 5.4 (4.3, 6.6) | 4.0 (2.2, 6.6) | 1.0 (0.4, 2.1) |
| Rh type |  |  |  |  |  |  |  |
|  | Rh positive | 16.1 (13.6, 18.7) | 1.6 (0.6, 3.4) | 1.9 (1.1, 3.1) | 4.9 (4.1, 5.8) | 4.4 (2.9, 6.4) | 1.3 (0.7, 2.4) |
|  | Rh negative | 13.4 (8.9, 19.0) | 0.9 (0.0, 3.9) | 1.7 (0.3, 5.1) | 1.6 (0.5, 3.7) | 3.1 (0.9, 7.7) | 2.1 (0.9, 4.1) |

**Figure 2.**
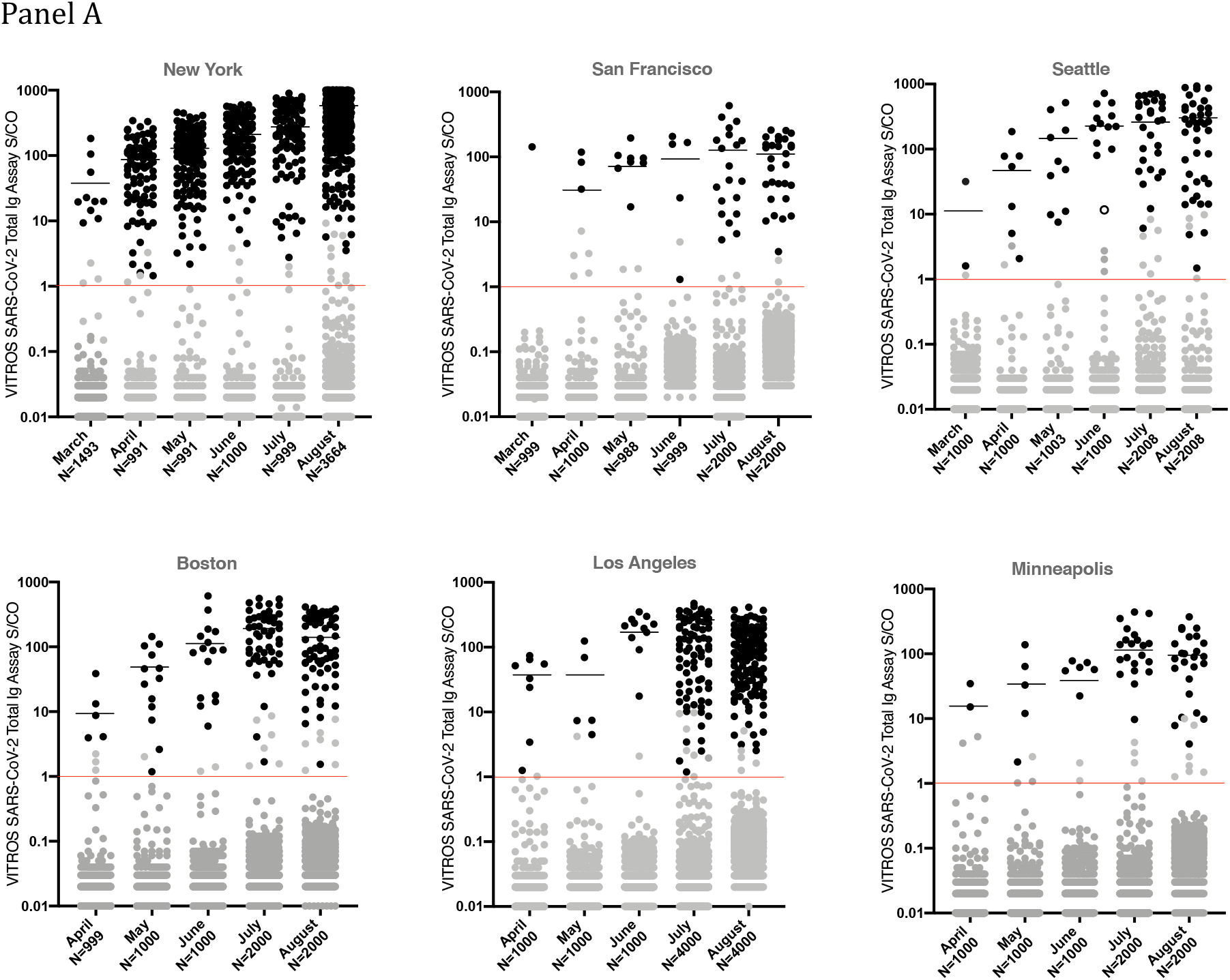
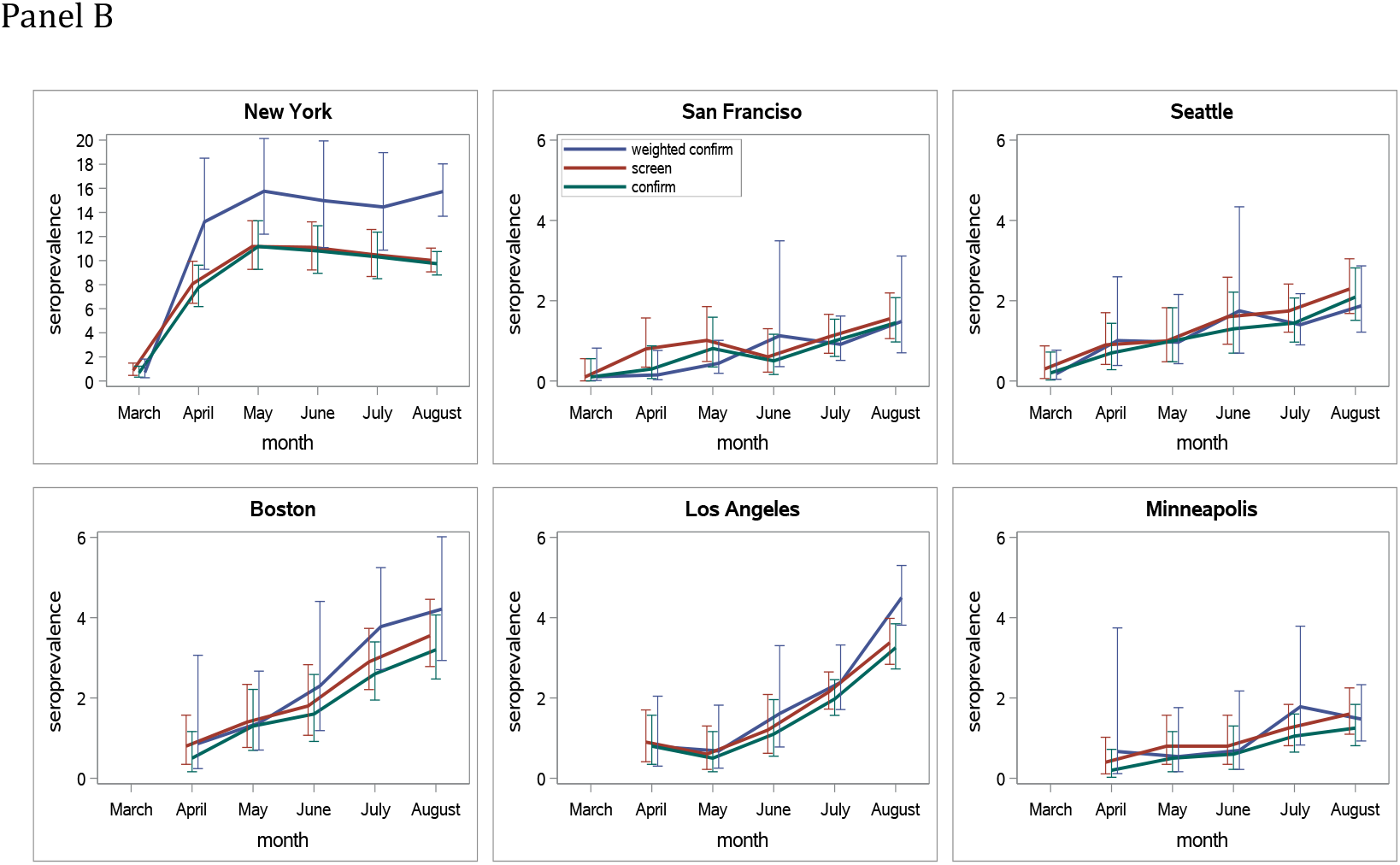
Monthly distribution of all Ortho Vitros SARS-CoV-2 Total Immunoglobulin (Ig) assay values (panel A) and unadjusted and weighted cumulative seroprevalence (panel B), six U.S. metropolitan regions, March–August 2020. Panel A: Red lines indicate the Vitros CoV2T signal to cutoff value for reactivity (S/CO ratio = 1.0; Log_10_ S/CO ratio = 0). Black symbols indicate confirmed reactive samples based on the study algorithm and black lines indicate the mean signal intensity of the Vitros CoV2T reactive donation samples for each region for each month of the study. Grey symbols above the Vitros CoV2T cutoff threshold indicate samples that were reactive by the Vitros CoV2T screening assay but which did not confirm using the study algorithm. Grey symbols below the red line indicate samples that were nonreactive on the Vitros CoV2T assay. The open black symbol (Seattle panel, June column) indicates the only sample with Vitros CoV2T S/CO >10 which did not confirm. N= Number of sampled donations for each month. Panel B: Screened and confirmed, and confirmed restricted to ZIP code of residence seroprevelence for each region.

In **Figure 2, Panel B**, the screening and confirmed seroprevalence data are presented over time for each DCR. A high proportion of screen-reactive donations confirmed, particularly in later months as seroprevalence increased; in July and August, 81-96% of specimens that screened reactive for anti-S antibodies by Vitros CoV2T were also reactive for anti-NC antibodies by Elecsys CoV2T.

Median weighted confirmed seroprevalence was 1.3 times higher than unweighted confirmed seroprevalences (IQR 1.02-1.44).

### Demographic, blood group and donation status associations with weighted seroprevalence estimates

The confirmed, weighted seroprevalence estimates by donor demographic subcategories (sex, age, race/ethnicity), and by blood groups (ABO and Rh) presented in **Table 2** were restricted to August as the most recent findings in this study. For New York, Los Angeles and Boston, sites with sufficient donations from racial and ethnic minority donors for meaningful comparison, seroprevalence was higher among younger age groups and among non-Hispanic Blacks and Hispanics than non-hispanic Whites. In New York in August, the seroprevalence among Hispanics was 28.6%, among non-Hispanic Blacks was 16.0% and among non-Hispanic Whites was 8.4%.

In a logistic regression model that included results from all regions and months, seroprevalence was associated with younger age (p<0.0001): compared to persons aged 50-64 years, persons aged 16-29 years had 1.31 (CI 1.1-1.6) times the odds of being seropositive. Both non-Hispanic Blacks (Odds ratio [OR] 2.2, 95% confidence interval [CI] 1.6-2.9) and Hispanics (OR 2.6, CI 2.2-3.1) had greater odds of being seropositive than non-Hispanic Whites **(Table 3)**. Gender and blood types were not significantly associated with seroprevalence. First-time donors had increased seroprevalence compared to repeat donors (OR 2.2, CI: 1.6-3.2)). In the four regions where donors in July and August were universally tested for SARS-CoV-2 antibodies, first-time donors had 2.2 (CI 1.8-2.6) times the odds of being seropositive compared to repeat donors. In the two regions where blood donors were not being offered antibody testing, first time donors had only 1.2 (CI 1.0-1.5) times the odds of repeat donors.

**Table 3.**
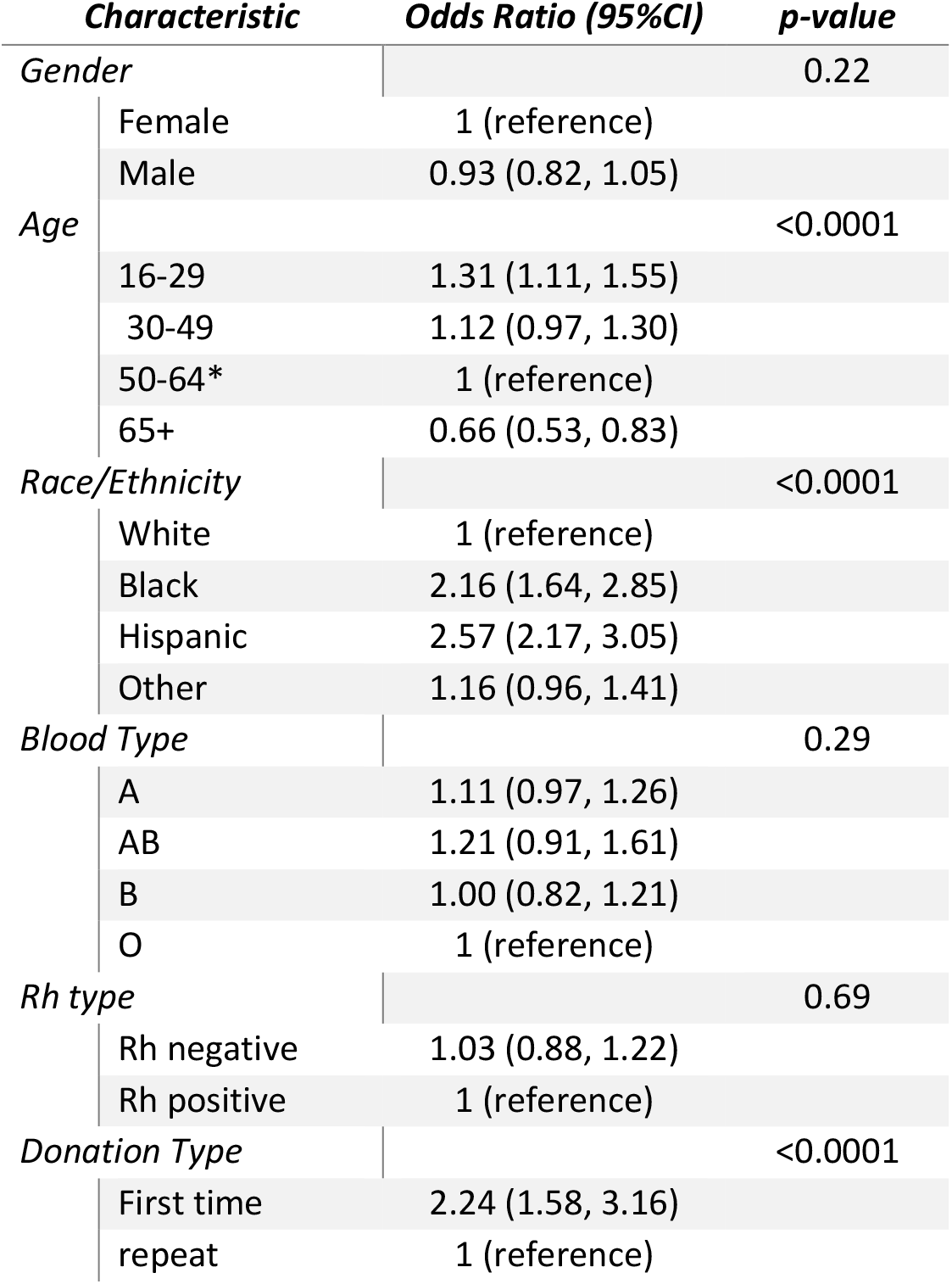
Factors associated with SARS-CoV-2 seropositivity in blood donors, six U.S. metropolitan regions, March–August 2020. 95% CI = 95% confidence interval. Rh = rhesus factor. *Reference 50-64 as the age group with highest frequency of donations

### Comparison of monthly seroprevalence (as calculated from donor serosurveillance) with reported COVID-19 case rates

For each region, the monthly confirmed, weighted seroprevalence was juxtaposed with the weekly and cumulative COVID-19 case counts. Seroprevalence and cumulative COVID-19 case rates increased in all regions from March/April through August **(Figure 3)**. New York reported the highest seroprevalence, increasing from 0.7% in March to 13.2% in April, corresponding with the sharp rise in reported New York COVID-19 cases. Coincident with a decrease in daily reported cases from May through July, the seroprevalence in New York stabilized at ∼15-16% during this time, with smaller increases in other regions. The cumulative case incidence for Boston and Los Angeles in July was similar to the cumulative case incidence for New York in April (∼2,000 cumulative reported cases per 100,000 population), but seroprevalence for Boston and Los Angeles remained substantially lower than New York.

**Figure 3.**
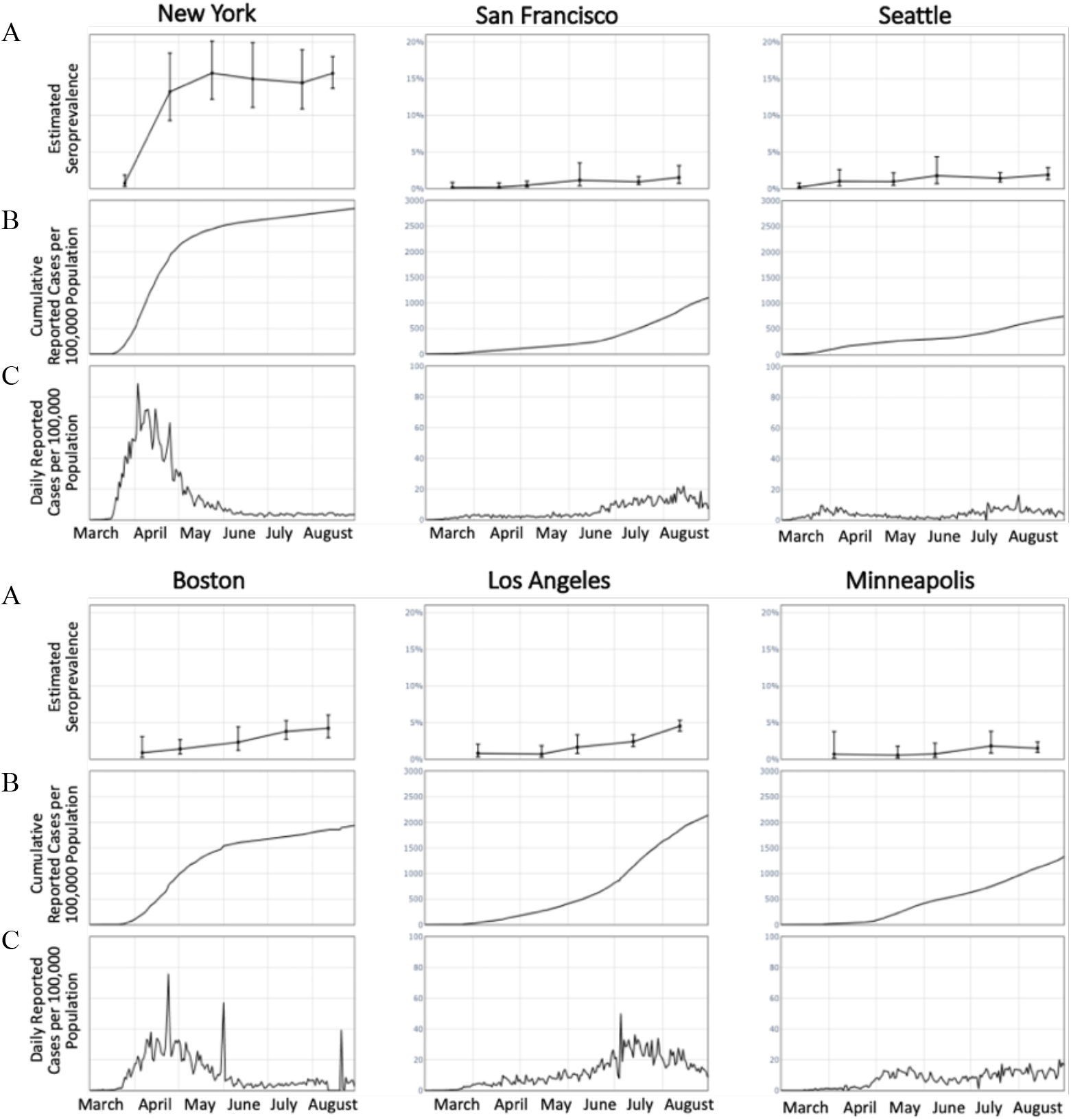
Weighted confirmed SARS-CoV-2 seroprevalence derived from blood donors (A), reported COVID-19 case rates per 100,000 population* (B), and daily COVID-19 case rates per 100,000* (C) in six U.S. metropolitan regions, March–August 2020. *Reported COVID-19 cases reported to CDC.

The number of estimated cumulative infections, based on the adjusted donor seroprevalence and population sizes, was larger than the number of cumulative reported infections for all regions **(Table 4)**. However, the ratio varied by region and over time. For all cities except New York, much higher numbers of estimated infections per reported case occurred in the first month of blood donor screening compared with later months. The highest reported ratio of estimated infections to reported cases occurred in Minneapolis in April (42 infections per reported case). By August 2020, all regions other than New York had 1.6-3.2 estimated infections per reported case. During May through August, New York had the highest number of estimated infections per reported case (5.3-6.4 infections per reported case).

**Table 4.**
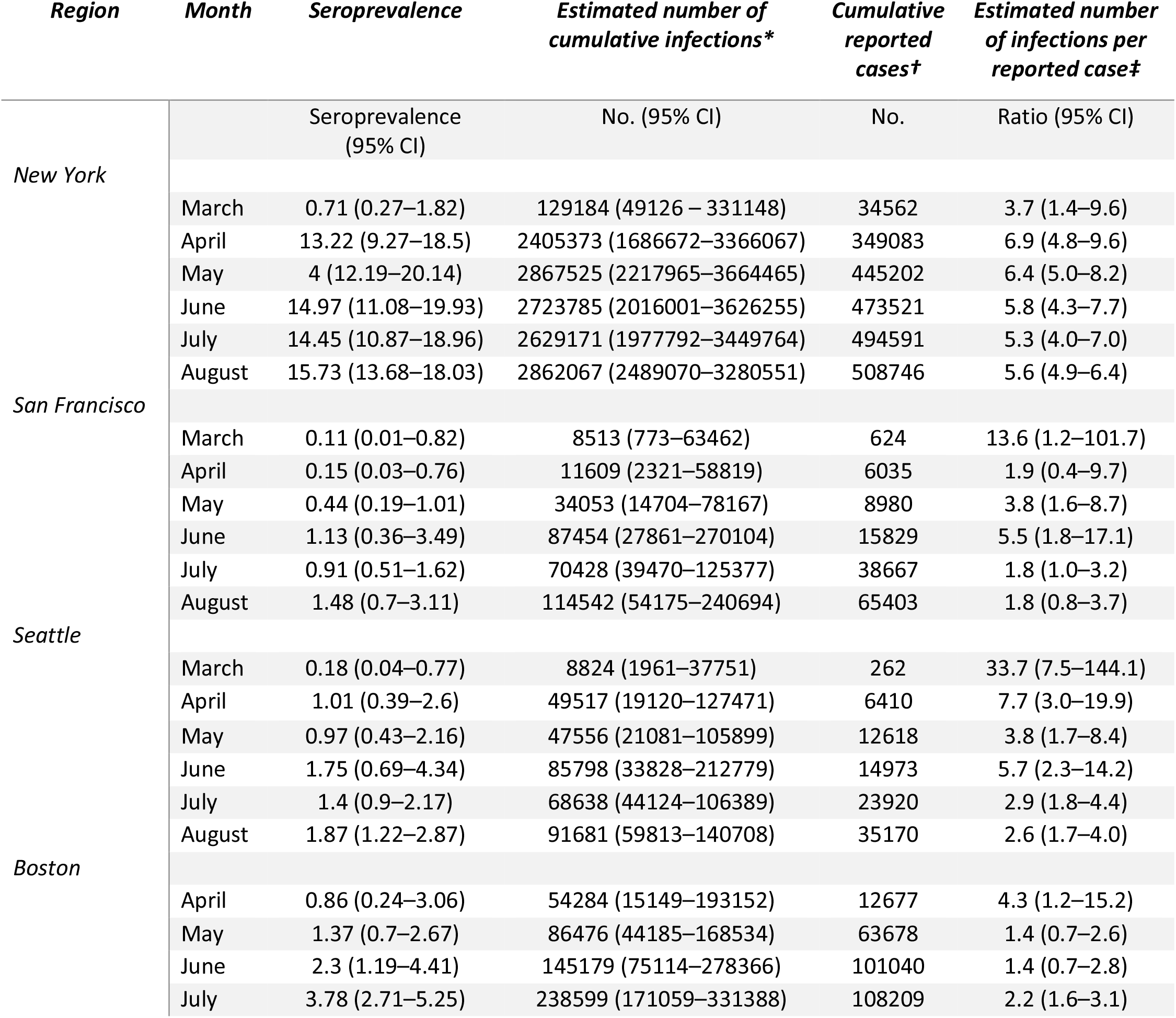
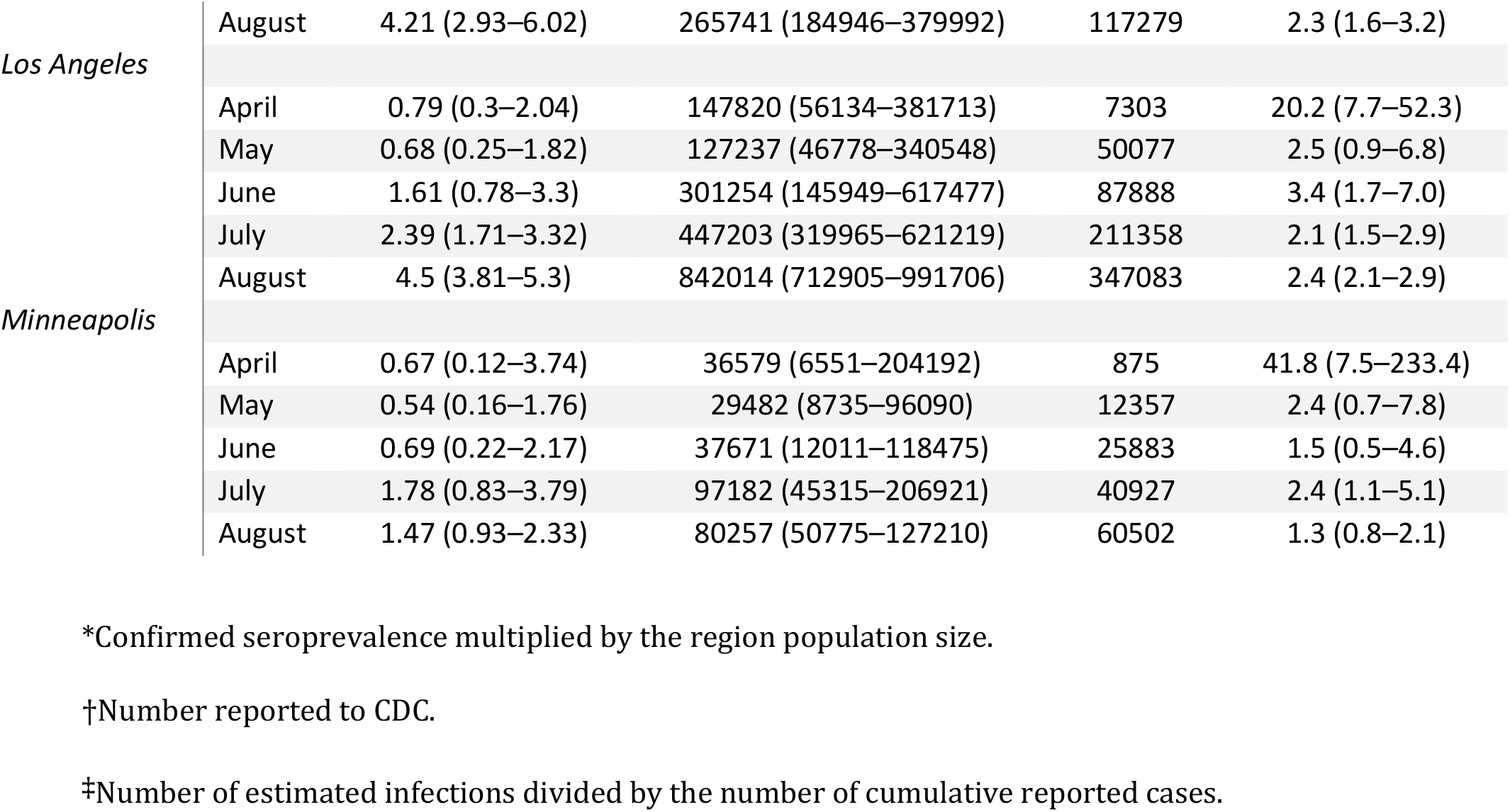
Monthly seroprevalence, estimated number of cumulative infections, cumulative reported COVID-19 cases, and the estimated number of cumulative infections per reported case, six U.S. metropolitan regions, March–August 2020. 95% CI = 95% confidence interval

## DISCUSSION

Use of blood donor populations with broad national representativeness provides a surveillance tool to monitor seroprevalence and to impute infection rates within communities, track outbreaks, and potentially correlate evolving infection rates with pandemic mitigation measures.

Critical to the success of serosurveillance programs is the choice of SARS-CoV-2 antibody assays and development and validation of supplemental testing algorithms. Antibody persistence or waning has been shown to be assay-dependent (22), so it is essential to select assays demonstrating durable antibody reactivity to accurately estimate cumulative incidence based on serial cross-sectional seroprevalence data. Also important is the assay’s ability to sensitively detect antibodies following asymptomatic and mildly symptomatic infections, which may produce weak systemic antibody responses (23).

The Vitros CoV2T and Elecsys CoV2T assays employed in this study satisfy many of these criteria for serosurveillance assays: They have stable S/CO values over at least 4-5 months following seroconversion (20, 24) and have wide dynamic ranges, enabling implementation of a screening assay S/CO threshold-based supplementary testing algorithm. By demonstrating that >99% of specimens screened with Vitros CoV2T that had S/CO≥10 were also reactive by the Elecsys CoV2T or RVPN, we were able to adopt a robust and lower-cost testing algorithm, limiting supplemental testing to screened specimens with S/CO 1-10. This algorithm is now being employed by CDC’s nationwide seroprevalence blood donor study. To differentiate natural-infection-induced and vaccine-induced seropositivity, the nationwide study is testing all anti-S-reactive specimens with an NC-based assay beginning in January 2021 (25).

Higher seroprevalence was observed in non-Hispanic Blacks and Hispanics than in non-Hispanic Whites in most regions but was particularly notable in New York. These racial seroprevalence disparities are consistent with other reports (15, 26), potentially because racial and ethnic minority groups experience inequities in access to health care, quality housing, ability to work from home, and reliable transportation (27). Increased risk for infection has been associated with younger age, possibly related to lack of adherence to mitigation measures (15, 28). Future analyses will include comparing region-, age-, and race/ethnicity-specific seroprevalence rates to the number of demographic group-specific cumulative reported cases.

In this study, seroprevalence trends were consistent with the pattern of cumulative reported COVID-19 cases. For most regions, the ratio of estimated infections to reported cases was higher during March-April 2020 than in subsequent months. This suggests that underreporting of COVID-19 cases to CDC was more severe during the earliest months of the pandemic. Lack of available testing and avoiding medical care to obtain testing because of COVID-19-related concerns might also have contributed (29). From May through August, the calculated seroprevalence predicted 1.6-3.2 SARS-CoV-2 infections per cumulative case reported to CDC for all regions except New York, which predicted 5.3-6.4 infections per reported case.

Compared to the other large seroprevalence survey conducted by CDC using commercial lab specimens, this study generally showed lower seroprevalence estimates (30). A national seroprevalence study of dialysis patients with blood specimens collected during July 2020 also reported generally higher seroprevalence estimates (31). Differences in the geographic distribution of participants, serology assays used, and assumptions made when extrapolating seroprevalence estimates to the general population may explain these differences. Several local seroprevalence studies conducted in regions similar to the six regions in this study have calculated similar or higher seroprevalence estimates (32-34). However, many of these collected specimens were from healthcare workers or hospitalized patients, who may be at higher risk of SARS-CoV-2 infection.

This study could have underestimated seroprevalence for several reasons. First, blood donors may represent a population less likely to be exposed to SARS-CoV-2 than the general population (35). Also, blood donors tend to be in better health than the general population and recruitment practices and eligibility criteria for blood donations may bias the donor sample toward lower-risk individuals; this may explain the lower rates of antibody positivity in repeat donors (who provide >80% of donations) compared to first time donors. Secondly, many higher-risk populations cannot or do not donate, including persons who are acutely febrile or ill, children aged <16 years, and institutionalized persons such as those residing in nursing homes or prison. Third, compared to the general population, relatively few ethnic and racial minorities donate, and these groups are at increased risk for SARS-CoV-2 infection; this bias is partially compensated for because our results were adjusted by weighting for race/ethnicity. Fourth, there is growing evidence that approximately 5-10% of infected persons do not seroconvert (23). We did not adjust our results to account for such “serosilent” infections.

Our results may overestimate seroprevalence because of implementation of SARS-CoV-2 antibody screening of all blood donations by some blood collection organizations in the summer of 2020. These blood centers publicly advertised availability of this screening which could have led to test seeking by prospective donors with increased concern over exposure to the virus. However, our analysis of relative seroprevalence before and after implementation of such “universal screening” in first time donors, who give 15-20% of total donations, indicates that although the odds ratio was greater for first-time donors, the impact of such test seeking was small relative to the expanding pandemic. Finally, there was no formal process for randomization, however no bias was seen in comparison of monthly samples with monthly donations (Supplemental Table 1).

Building on the approach developed in the RESPONSE seroprevalence study, in July 2020 the U.S. CDC funded a nationwide blood donor seroprevalence program that expanded this surveillance program from six regions for 6 months to >60 U.S. regions with monthly collections of 2,000-6,000 samples per region from July 2020 to December 2021 (Supplemental Figure 2). Similar to RESPONSE, changes in overall, geographic region-, age-, sex-, and race/ethnicity-specific SARS-CoV-2 seroprevalence will be calculated monthly over the course of the study and compared with clinical cases, deaths, and community serosurvey data.

In conclusion, serial serosurveillance studies of SARS-CoV-2 using blood donor populations, which are now being implemented in many countries (36), provide a powerful adjunct to standard public health case reporting. Although serosurveillance data from asymptomatic blood donors may lag behind viral transmission and case reporting by up to several weeks, if appropriately designed, executed, analyzed, and interpreted, these studies will provide urgently needed data to inform our understanding of the epidemiology and effectiveness of responses to this unprecedented pandemic.

## Data Availability

The data is not being made publicly available for privacy reasons related to federal regulations but a limited dataset may be made available upon request after formal review by NHLBI and CDC's privacy office before release.

## Acknowledgments

The NHLBI REDS Epidemiology, Surveillance and Preparedness of the Novel SARS-CoV-2 (RESPONSE) study is the responsibility of the following persons: Vitalant Research Institute M.P. Busch, P.J. Norris, and M. Stone, Vitalant Research Institute, San Francisco, CA, Data coordinating center; S.M. Mathew, Westat, Rockville, MD; Blood Collection Organizations: S. Stramer, American Red Cross (ARC), Gaithersburg, MD, D. Kessler, New York Blood Center (NYBC), New York, NY, B.A. Konkle, Blood Works Northwest, Seattle, WA, B. Custer, Vitalant Research Institute, San Francisco, CA; Publications Committee Chairman: P.M. Ness, Johns Hopkins University, Baltimore, MD; Steering Committee Chairpersons: S.H. Kleinman, University of British Columbia, Victoria, BC, Canada, C.D. Josephson, Emory University, Atlanta, GA, National Heart, Lung, and Blood Institute, National Institutes of Health, S.A. Glynn and K. Malkin.

The authors thank C. Cassetti, National Institute of Allergies and Infectious Diseases (NIAID) and J. Jones, S. Gerber, M. Patton, F. Havers and S. Basavaraju of the Centers for Disease Control and Prevention (CDC) for their technical support, as well as A.E. Williams and S. Anderson of the U.S. Food and Drug Administration (FDA), J. Haynes from ARC for their contribution of data from the Transfusion-Transmissible Infections Monitoring System (TTIMS) and L. McCain, A. Hui, C. Samuels, H. Tanner and Z. Kaidarova of Vitalant Research Institute for their technical assistance.

