## Supplemental Material for "Use of U.S. Blood Donors for National Serosurveillance of SARS-CoV-2 Antibodies: Basis for an Expanded National Donor Serosurveillance Program"

Supplemental Table 1a. New York; Comparison of monthly samples with monthly donations

| **Variable** | **March** | **April** | **May** | **June** | **July** | **August** |
| --- | --- | --- | --- | --- | --- | --- |
|  | **Donations/Samples** | **Donations/Samples** | **Donations/Samples** | **Donations/Samples** | **Donations/Samples** | **Donations/Samples** |
| **Total** | 28432 / 1493 | 15505 / 991 | 20114 / 985 | 25565 / 1000 | 21003 / 999 | 22103 / 3664 |
| **Gender** | p<0.001 | p=0.01 | p<0.001 | p=0.30 | p=0.51 | p<0.001 |
| Female | 44.9 / 54.5 | 46.4 / 50.3 | 49.3 / 60.6 | 48.1 / 46.4 | 45.4 / 46.4 | 43.9 / 40.9 |
| Male | 55.1 / 45.5 | 53.6 / 49.7 | 50.7 / 39.4 | 51.9 / 53.6 | 54.6 / 53.6 | 56.1 / 59.1 |
| **Age** | p<0.001 | p=0.39 | p<0.001 | p=0.003 | p=0.17 | p=0.002 |
| 16-29 | 22.7 / 20.2 | 15.8 / 15.3 | 15.6 / 19.7 | 18.3 / 21.4 | 16.7 / 18.3 | 21.0 / 22.6 |
| 30-49 | 29.3 / 33.8 | 30.8 / 30.7 | 30.2 / 36.8 | 32.5 / 35.3 | 29.0 / 30.1 | 28.2 / 27.9 |
| 50-64 | 37.1 / 34.5 | 41.6 / 40.4 | 41.1 / 31.7 | 36.2 / 31.9 | 39.5 / 36.2 | 36.5 / 34.0 |
| 65+ | 10.9 / 11.5 | 11.9 / 13.6 | 13.1 / 11.9 | 12.9 / 11.4 | 14.8 / 15.3 | 14.3 / 15.6 |
| **Race/Ethnicity** | p<0.001 | p=0.38 | p=0.43 | p<0.001 | p=0.37 | p<0.001 |
| White | 77.2 / 84.7 | 83.6 / 81.8 | 80.9 / 79.3 | 76.8 / 70.2 | 77.5 / 76.2 | 75.9 / 74.7 |
| Black | 3.9 / 1.7 | 2.7 / 2.6 | 3.2 / 3.9 | 3.9 / 6.3 | 3.8 / 3.6 | 4.4 / 4.3 |
| Hispanic | 9.6 / 5.7 | 6.7 / 7.7 | 8.7 / 8.7 | 9.5 / 8.1 | 9.1 / 8.9 | 9.5 / 8.6 |
| Other | 9.4 / 8.0 | 6.9 / 7.9 | 7.2 / 8.1 | 9.8 / 15.4 | 9.7 / 11.3 | 10.2 / 12.4 |
| **Blood group** | p=0.10 | p=0.20 | p=0.02 | p=0.03 | p=0.88 | p=0.49 |
| A | 50.2 / 47.5 | 48.8 / 52.2 | 51.0 / 46.2 | 51.4 / 47.0 | 51.7 / 51.0 | 50.9 / 50.5 |
| AB | 32.9 / 35.8 | 32.7 / 30.5 | 33.7 / 37.4 | 32.3 / 34.1 | 32.2 / 32.1 | 33.0 / 32.6 |
| B | 12.8 / 12.4 | 13.0 / 12.1 | 11.2 / 11.7 | 11.9 / 14.2 | 12.3 / 13.1 | 12.1 / 12.3 |
| O | 4.1 / 4.2 | 5.5 / 5.1 | 4.0 / 4.7 | 4.4 / 4.5 | 3.8 / 3.7 | 4.1 / 4.6 |
| **Rh Type** | p=0.29 | p=0.010 | p=0.86 | p=0.01 | p=0.24 | p<0.001 |
| Rh positive | 83.8 / 82.6 | 83.3 / 80.1 | 82.5 / 82.2 | 83.4 / 86.2 | 83.2 / 84.5 | 83.7 / 85.8 |
| Rh negative | 16.2 / 17.2 | 16.7 / 19.8 | 17.5 / 17.7 | 16.6 / 13.6 | 16.8 / 15.4 | 16.3 / 14.2 |
| **Donor Status** | p<0.001 | p<0.001 | p<0.001 | p=0.08 | p=0.003 | p=0.13 |
| First time | 15.8 / 23.4 | 22.0 / 16.5 | 14.7 / 20.8 | 16.2 / 14.2 | 10.2 / 7.3 | 10.8 / 11.5 |
| Repeat | 84.2 / 76.6 | 78.0 / 83.5 | 85.3 / 79.2 | 83.8 / 85.8 | 89.8 / 92.7 | 89.2 / 88.5 |

Supplemental Table 1b. San Francisco; Comparison of monthly samples with monthly donations

| **Variable** | **March** | **April** | **May** | **June** | **July** | **August** |
| --- | --- | --- | --- | --- | --- | --- |
|  | **donations/samples** | **donations/samples** | **donations/samples** | **donations/samples** | **donations/samples** | **donations/samples** |
| **Total** | 4342 / 999 | 3840 / 1000 | 3792 / 988 | 6144 / 999 | 5670 / 2000 | 4970 / 2000 |
| **Gender** | p=0.35 | p=0.54 | p=0.55 | p=0.009 | p=0.62 | p=0.08 |
| Female | 47.7 / 49.1 | 53.7 / 52.8 | 53.2 / 52.2 | 51.4 / 47.2 | 52.8 / 53.4 | 51.1 / 49.1 |
| Male | 52.3 / 50.9 | 46.3 / 47.2 | 46.8 / 47.8 | 48.6 / 52.8 | 47.2 / 46.6 | 48.9 / 50.9 |
| **Age** | p<0.001 | p=0.12 | p=0.05 | p<0.001 | p=0.19 | p=0.65 |
| 16-29 | 15.0 / 17.0 | 12.2 / 11.3 | 11.5 / 9.0 | 11.8 / 7.4 | 12.1 / 11.6 | 12.7 / 13.2 |
| 30-49 | 29.8 / 37.7 | 32.2 / 34.2 | 28.3 / 29.9 | 31.1 / 29.5 | 33.4 / 32.0 | 29.3 / 30.2 |
| 50-64 | 34.9 / 29.0 | 37.7 / 34.8 | 39.7 / 39.0 | 36.4 / 39.7 | 36.0 / 36.4 | 37.4 / 36.8 |
| 65+ | 20.4 / 16.2 | 17.9 / 19.6 | 20.5 / 22.2 | 20.8 / 23.3 | 18.5 / 20.1 | 20.6 / 19.9 |
| **Race/Ethnicity** | p=0.89 | p=0.84 | p=0.95 | p=0.49 | p=0.18 | p=0.88 |
| White | 69.7 / 69.8 | 71.5 / 71.7 | 73.5 / 73.4 | 72.7 / 72.6 | 69.8 / 71.6 | 70.7 / 70.0 |
| Black | 1.9 / 1.6 | 1.1 / 0.8 | 1.3 / 1.4 | 1.2 / 1.7 | 1.5 / 1.7 | 1.3 / 1.3 |
| Hispanic | 9.1 / 9.5 | 7.4 / 7.2 | 7.1 / 7.5 | 7.5 / 7.6 | 8.6 / 8.6 | 9.1 / 9.2 |
| Other | 19.4 / 19.1 | 19.9 / 20.3 | 18.0 / 17.7 | 18.7 / 18.1 | 20.1 / 18.2 | 18.9 / 19.6 |
| **Blood group** | p<0.001 | p=0.43 | p=0.48 | p=0.39 | p=0.52 | p=0.54 |
| A | 48.8 / 48.6 | 45.8 / 45.4 | 49.0 / 48.0 | 49.6 / 50.3 | 48.3 / 47.8 | 49.4 / 49.0 |
| AB | 32.6 / 33.9 | 34.0 / 32.9 | 32.7 / 34.8 | 34.2 / 32.5 | 33.6 / 35.1 | 34.4 / 35.6 |
| B | 12.3 / 14.3 | 12.6 / 12.7 | 12.2 / 11.7 | 11.7 / 13.1 | 13.1 / 12.5 | 11.3 / 11.2 |
| O | 6.3 / 3.1 | 7.6 / 8.9 | 6.1 / 5.5 | 4.5 / 4.1 | 5.0 / 4.8 | 4.8 / 4.3 |
| **Rh Type** | p=0.41 | p=0.65 | p=0.71 | p=0.81 | p=0.34 | p=0.05 |
| Rh positive | 83.4 / 84.4 | 83.4 / 82.8 | 82.9 / 83.3 | 80.9 / 80.6 | 84.4 / 83.6 | 83.2 / 84.9 |
| Rh negative | 16.6 / 15.6 | 16.6 / 17.1 | 17.1 / 16.7 | 19.1 / 19.4 | 15.6 / 16.4 | 16.8 / 15.2 |
| **Donor status** | p=0.59 | p<0.001 | p<0.001 | p<0.001 | p<0.001 | p<0.001 |
| First time | 21.2 / 20.5 | 38.5 / 18.1 | 33.0 / 19.1 | 25.4 / 12.4 | 32.0 / 21.4 | 22.9 / 15.8 |
| Repeat | 78.8 / 79.5 | 61.5 / 81.9 | 67.0 / 80.9 | 74.6 / 87.6 | 68.0 / 78.7 | 77.1 / 84.3 |

Supplemental Table 1c. Seattle; Comparison of monthly samples with monthly donations

| **Variable** | **March** | **April** | **May** | **June** | **July** | **August** |
| --- | --- | --- | --- | --- | --- | --- |
|  | **donations/samples** | **donations/samples** | **donations/samples** | **donations/samples** | **donations/samples** | **donations/samples** |
| **Total** | 13972 / 1000 | 9831 / 1000 | 13304 / 1003 | 12053 / 1000 | 14041 / 2008 | 13008 / 2008 |
| **Gender** | p=0.18 | p=0.28 | p=0.22 | p=0.10 | p=0.23 | p=0.76 |
| Female | 56.2 / 54.1 | 57.7 / 59.4 | 57.2 / 55.2 | 56.1 / 53.5 | 57.0 / 55.7 | 56.2 / 55.8 |
| Male | 43.8 / 45.9 | 42.3 / 40.6 | 42.8 / 44.8 | 43.9 / 46.5 | 43.0 / 44.3 | 43.8 / 44.2 |
| **Age** | p=0.002 | p=0.63 | p<0.001 | p=0.08 | p=0.03 | p=0.008 |
| 16-29 | 15.3 / 12.6 | 14.0 / 13.3 | 13.6 / 11.4 | 14.7 / 14.2 | 14.4 / 13.7 | 14.1 / 12.9 |
| 30-49 | 34.4 / 31.2 | 35.4 / 37.2 | 34.3 / 31.8 | 33.2 / 29.9 | 33.8 / 31.9 | 34.6 / 33.5 |
| 50-64 | 31.3 / 35.1 | 32.8 / 32.5 | 33.3 / 33.0 | 31.9 / 33.4 | 32.2 / 32.4 | 31.6 / 31.0 |
| 65+ | 19.1 / 21.1 | 17.9 / 17.0 | 18.7 / 23.8 | 20.3 / 22.5 | 19.5 / 22.0 | 19.6 / 22.6 |
| **Race/Ethnicity** | p=0.19 | p=0.90 | p=0.13 | p=0.13 | p<0.001 | p=0.01 |
| White | 82.3 / 84.3 | 81.0 / 81.1 | 81.3 / 83.8 | 80.6 / 83.4 | 80.9 / 85.5 | 80.9 / 83.2 |
| Black | 0.7 / 0.5 | 0.8 / 0.9 | 0.9 / 0.6 | 1.0 / 0.7 | 1.0 / 0.5 | 0.9 / 1.0 |
| Hispanic | 2.3 / 2.6 | 1.9 / 2.1 | 2.2 / 1.5 | 2.5 / 1.9 | 2.6 / 2.4 | 2.6 / 2.9 |
| Other | 14.7 / 12.6 | 16.3 / 15.9 | 15.6 / 14.1 | 16.0 / 14.0 | 15.5 / 11.6 | 15.6 / 12.9 |
| **Blood group** | p=0.26 | p=0.44 | p=0.41 | p=0.24 | p<0.001 | p=0.18 |
| A | 49.1 / 51.9 | 49.1 / 48.2 | 52.3 / 51.1 | 50.7 / 53.2 | 51.1 / 53.9 | 49.7 / 51.9 |
| AB | 36.3 / 34.8 | 34.8 / 33.8 | 34.0 / 36.1 | 36.1 / 34.0 | 35.6 / 37.4 | 34.6 / 33.0 |
| B | 10.3 / 9.9 | 10.8 / 12.2 | 9.7 / 9.5 | 9.4 / 8.5 | 9.6 / 8.3 | 12.1 / 11.2 |
| O | 4.2 / 3.4 | 5.3 / 5.8 | 4.0 / 3.3 | 3.8 / 4.3 | 3.7 / 0.4 | 3.7 / 3.7 |
| **Rh Type** | p=0.04 | p=0.52 | p=0.61 | p=0.36 | p=0.04 | p=0.24 |
| Rh positive | 80.3 / 77.7 | 79.9 / 80.7 | 78.9 / 78.3 | 80.8 / 79.7 | 79.1 / 77.1 | 80.7 / 81.6 |
| Rh negative | 19.7 / 22.3 | 20.1 / 19.3 | 21.1 / 21.7 | 19.2 / 20.3 | 20.9 / 22.8 | 19.3 / 18.2 |
| **Donor status** | p<0.001 | p=0.19 | p=0.78 | p<0.001 | p<0.001 | p=0.21 |
| First time | 26.8 / 17.2 | 33.4 / 31.5 | 23.7 / 23.3 | 14.7 / 9.9 | 13.7 / 7.8 | 17.1 / 16.0 |
| Repeat | 73.2 / 82.8 | 66.6 / 68.5 | 76.3 / 76.7 | 85.3 / 90.1 | 86.3 / 92.2 | 82.9 / 84.0 |
|  | 78.8 / 79.5 | 61.5 / 81.9 | 67.0 / 80.9 | 74.6 / 87.6 | 68.0 / 78.7 | 77.1 / 84.3 |

Supplemental Table 1d. Los Angeles; Comparison of monthly samples with monthly donations

| **Variable** | **March** | **April** | **May** | **June** | **July** | **August** |
| --- | --- | --- | --- | --- | --- | --- |
|  | **donations/samples** | **donations/samples** | **donations/samples** | **donations/samples** | **donations/samples** | **donations/samples** |
| **Total** |  | 17242 / 1000 | 21435 / 1000 | 25647 / 1000 | 26738 / 4000 | 23630 / 4000 |
| **Gender** |  | p=0.47 | p=0.17 | p=0.77 | p=0.91 | p=0.40 |
| Female |  | 55.5 / 54.4 | 54.3 / 52.1 | 54.3 / 53.8 | 55.9 / 55.8 | 53.6 / 53.0 |
| Male |  | 44.5 / 45.6 | 45.7 / 47.9 | 45.7 / 46.2 | 44.1 / 44.2 | 46.4 / 47.1 |
| **Age** |  | p=0.27 | p=0.12 | p=0.30 | p=0.05 | p=0.71 |
| 16-29 |  | 15.6 / 15.5 | 15.5 / 13.1 | 17.0 / 18.2 | 16.7 / 16.7 | 16.3 / 16.6 |
| 30-49 |  | 34.2 / 32.1 | 31.5 / 30.8 | 35.8 / 33.0 | 38.9 / 40.9 | 35.7 / 34.9 |
| 50-64 |  | 36.3 / 39.2 | 37.2 / 39.8 | 33.3 / 34.7 | 32.3 / 30.9 | 34.5 / 35.0 |
| 65+ |  | 13.9 / 13.2 | 15.8 / 16.3 | 13.9 / 14.1 | 12.1 / 11.5 | 13.5 / 13.6 |
| **Race/Ethnicity** |  | p=0.002 | p=0.08 | p=0.04 | p=0.97 | p=0.02 |
| White |  | 66.6 / 62.0 | 63.6 / 61.4 | 62.1 / 65.0 | 60.1 / 60.3 | 61.0 / 59.1 |
| Black |  | 1.9 / 2.6 | 2.3 / 1.6 | 2.2 / 2.8 | 2.0 / 2.0 | 2.1 / 2.5 |
| Hispanic |  | 17.7 / 18.2 | 19.1 / 19.9 | 19.8 / 16.7 | 21.1 / 21.2 | 20.7 / 22.3 |
| Other |  | 13.7 / 17.2 | 14.9 / 17.1 | 16.0 / 15.5 | 16.8 / 16.6 | 16.2 / 16.1 |
| **Blood group** |  | p=0.32 | p=0.05 | p=0.16 | p=0.15 | p=0.23 |
| A |  | 49.6 / 49.4 | 51.6 / 48.2 | 50.6 / 54.1 | 50.1 / 48.5 | 51.6 / 52.7 |
| AB |  | 33.4 / 31.6 | 32.6 / 36.3 | 32.9 / 30.9 | 32.8 / 33.5 | 32.2 / 30.9 |
| B |  | 11.2 / 12.6 | 11.8 / 11.0 | 12.1 / 11.2 | 12.2 / 13.2 | 12.0 / 11.9 |
| O |  | 5.8 / 6.4 | 3.9 / 4.5 | 4.5 / 3.8 | 4.8 / 4.8 | 4.2 / 4.6 |
| **Rh Type** |  | p=0.92 | p=0.61 | p=0.54 | p=0.24 | p=0.66 |
| Rh positive |  | 84.5 / 84.4 | 83.9 / 83.3 | 85.0 / 84.3 | 84.8 / 84.2 | 84.9 / 85.1 |
| Rh negative |  | 15.5 / 15.6 | 16.1 / 16.7 | 15.0 / 15.7 | 15.2 / 15.8 | 15.1 / 14.9 |
| **Donor status** |  | p<0.001 | p<0.001 | p<0.001 | p=0.58 | p=0.06 |
| First time |  | 41.7 / 23.9 | 28.4 / 13.7 | 28.2 / 20.0 | 33.1 / 33.5 | 25.9 / 27.2 |
| Repeat |  | 58.3 / 76.1 | 71.6 / 86.3 | 71.8 / 80.0 | 66.9 / 66.5 | 74.1 / 72.9 |

Supplemental Table 1e. Boston; Comparison of monthly samples with monthly donations

| **Variable** | **March** | **April** | **May** | **June** | **July** | **August** |
| --- | --- | --- | --- | --- | --- | --- |
|  | **donations/samples** | **donations/samples** | **donations/samples** | **donations/samples** | **donations/samples** | **donations/samples** |
| **Total** |  | 7344 / 999 | 8336 / 1000 | 10327 / 1000 | 10939 / 2000 | 10491 / 2000 |
| **Gender** |  | p=0.97 | p=0.15 | p=0.42 | p=0.42 | p=0.24 |
| Female |  | 53.8 / 53.8 | 52.5 / 54.8 | 53.0 / 51.7 | 55.8 / 56.8 | 52.5 / 51.3 |
| Male |  | 46.2 / 46.2 | 47.5 / 45.2 | 47.0 / 48.3 | 44.2 / 43.3 | 47.5 / 48.8 |
| **Age** |  | p<0.001 | p=0.10 | p=0.75 | p=0.68 | p=0.002 |
| 16-29 |  | 13.1 / 16.7 | 12.2 / 11.8 | 14.0 / 13.3 | 14.6 / 13.8 | 13.7 / 15.8 |
| 30-49 |  | 28.9 / 32.4 | 25.7 / 28.7 | 27.2 / 26.4 | 31.1 / 31.9 | 28.8 / 26.2 |
| 50-64 |  | 41.3 / 36.9 | 42.9 / 42.6 | 40.2 / 40.9 | 38.0 / 38.5 | 39.8 / 38.7 |
| 65+ |  | 16.7 / 13.9 | 19.2 / 16.9 | 18.5 / 19.4 | 16.2 / 15.9 | 17.7 / 19.4 |
| **Race/Ethnicity** |  | p=0.69 | p=0.62 | p=0.54 | p=0.78 | p=0.73 |
| White |  | 93.8 / 93.3 | 93.2 / 93.4 | 92.3 / 92.9 | 92.0 / 92.4 | 92.2 / 92.2 |
| Black |  | 0.7 / 0.8 | 1.0 / 1.3 | 1.2 / 0.7 | 1.1 / 1.1 | 0.9 / 1.1 |
| Hispanic |  | 1.7 / 2.2 | 2.0 / 2.0 | 2.1 / 1.9 | 2.1 / 2.2 | 2.1 / 2.4 |
| Other |  | 3.8 / 3.7 | 3.8 / 3.3 | 4.4 / 4.5 | 4.8 / 4.3 | 4.7 / 4.4 |
| **Blood group** |  | p=0.008 | p=0.68 | p=0.68 | p=0.43 | p=0.77 |
| A |  | 50.3 / 48.5 | 50.4 / 52.1 | 52.6 / 53.5 | 51.2 / 51.2 | 51.1 / 52.2 |
| AB |  | 33.4 / 33.3 | 34.6 / 33.3 | 32.2 / 32.0 | 33.2 / 34.2 | 33.1 / 32.4 |
| B |  | 11.4 / 11.0 | 11.0 / 10.4 | 11.0 / 11.0 | 11.1 / 10.1 | 11.3 / 11.2 |
| O |  | 4.8 / 7.1 | 4.0 / 4.2 | 4.3 / 3.5 | 4.5 / 4.6 | 4.4 / 4.1 |
| **Rh Type** |  | p=0.87 | p=0.82 | p=0.55 | p=0.96 | p=0.80 |
| Rh positive |  | 81.8 / 81.6 | 80.5 / 80.2 | 79.9 / 79.1 | 80.6 / 80.7 | 81.0 / 80.7 |
| Rh negative |  | 18.2 / 18.4 | 19.5 / 19.8 | 20.1 / 20.9 | 19.4 / 19.4 | 18.9 / 19.2 |
| **Donor status** |  | p<0.001 | p=0.03 | p<0.001 | p=0.78 | p=0.02 |
| First time |  | 33.4 / 27.5 | 24.0 / 26.9 | 18.4 / 10.4 | 23.1 / 23.4 | 18.6 / 20.7 |
| Repeat |  | 66.6 / 72.5 | 76.0 / 73.1 | 81.6 / 89.6 | 76.9 / 76.7 | 81.4 / 79.3 |

Supplemental Table 1f. Minneapolis; Comparison of monthly samples with monthly donations

| **Variable** |  | **April** | **May** | **June** | **July** | **August** |
| --- | --- | --- | --- | --- | --- | --- |
|  |  | **donations/samples** | **donations/samples** | **donations/samples** | **donations/samples** | **donations/samples** |
| **Total** |  | 15400 / 1000 | 16873 / 1000 | 21538 / 1000 | 22820 / 2000 | 21379 / 2000 |
| **Gender** |  | p=0.02 | p=0.69 | p=0.36 | p=0.70 | p=0.73 |
| Female |  | 57.6 / 53.9 | 55.6 / 56.2 | 56.6 / 58.0 | 59.3 / 59.7 | 58.0 / 58.4 |
| Male |  | 42.4 / 46.1 | 44.4 / 43.8 | 43.4 / 42.0 | 40.7 / 40.3 | 42.0 / 41.6 |
| **Age** |  | p<0.001 | p=0.03 | p=0.03 | p=0.94 | p=0.98 |
| 16-29 |  | 12.4 / 8.8 | 10.6 / 9.7 | 10.5 / 8.4 | 10.6 / 10.4 | 10.4 / 10.4 |
| 30-49 |  | 26.5 / 26.0 | 23.7 / 27.7 | 27.7 / 25.7 | 31.7 / 32.2 | 29.7 / 29.6 |
| 50-64 |  | 37.3 / 42.2 | 37.6 / 35.8 | 35.5 / 37.6 | 35.3 / 34.8 | 35.6 / 36.0 |
| 65+ |  | 23.8 / 23.0 | 28.1 / 26.8 | 26.3 / 28.3 | 22.5 / 22.6 | 24.4 / 24.1 |
| **Race/Ethnicity** |  | p=0.07 | p=0.65 | p=0.91 | p=0.19 | p=0.87 |
| White |  | 97.2 / 97.9 | 97.0 / 96.4 | 97.0 / 97.1 | 96.9 / 96.7 | 96.8 / 97.0 |
| Black |  | 0.3 / 0.5 | 0.4 / 0.4 | 0.3 / 0.3 | 0.3 / 0.4 | 0.4 / 0.3 |
| Hispanic |  | 0.8 / 0.8 | 0.9 / 0.9 | 0.8 / 0.6 | 0.8 / 0.5 | 0.9 / 0.8 |
| Other |  | 1.7 / 0.8 | 1.8 / 2.3 | 1.9 / 2.0 | 1.9 / 2.5 | 2.0 / 2.0 |
| **Blood group** |  | p=0.18 | p=0.61 | p=0.96 | p=0.38 | p=0.48 |
| A |  | 47.0 / 46.0 | 47.9 / 49.5 | 48.3 / 48.4 | 47.7 / 47.2 | 46.7 / 46.6 |
| AB |  | 38.1 / 39.9 | 36.8 / 34.8 | 36.5 / 36.4 | 37.0 / 36.9 | 38.0 / 37.2 |
| B |  | 10.4 / 8.8 | 10.6 / 11.1 | 10.4 / 10.2 | 10.7 / 11.8 | 10.8 / 11.8 |
| O |  | 4.5 / 5.3 | 4.6 / 4.6 | 4.7 / 5.0 | 4.6 / 4.1 | 4.5 / 4.4 |
| **Rh Type** |  | p=0.16 | p=0.45 | p=0.71 | p=0.40 | p=0.01 |
| Rh positive |  | 78.7 / 80.5 | 77.7 / 76.7 | 78.4 / 78.9 | 78.8 / 78.0 | 79.0 / 76.7 |
| Rh negative |  | 21.3 / 19.5 | 22.3 / 23.3 | 21.6 / 21.1 | 21.2 / 22.0 | 21.0 / 23.3 |
| **Donor status** |  | p<0.001 | p=0.03 | p<0.001 | p=0.26 | p=0.92 |
| First time |  | 24.2 / 17.6 | 14.3 / 11.9 | 13.5 / 8.7 | 21.5 / 20.5 | 17.1 / 17.2 |
| Repeat |  | 75.8 / 82.4 | 85.7 / 88.1 | 86.5 / 91.3 | 78.5 / 79.5 | 82.9 / 82.8 |

|  |  | | **Blood Center Samples** | | | **DCR weighted samples** | | | |
| --- | --- | --- | --- | --- | --- | --- | --- | --- | --- |
| **Center** | **Month** | **N** | | **Screen (95%CI)** | **Confirm (95%CI)** | | **N in DCR** | **DCR Screen (95%CI)** | **DCR Confirm (95%CI)** |
| **New York** |  |  | |  |  | |  |  |  |
|  | March | 1493 | | 0.87 (0.46, 1.48) | 0.67 (0.32, 1.23) | | 1331 | 1.30 (0.59, 2.84) | 0.71 (0.27, 1.82) |
|  | April | 991 | | 8.07 (6.45, 9.95) | 7.77 (6.18, 9.62) | | 874 | 13.26 (9.31, 18.53) | 13.22 (9.27, 18.50) |
|  | May | 985 | | 11.17 (9.27, 13.30) | 11.17 (9.27, 13.30) | | 854 | 15.76 (12.19, 20.14) | 15.76 (12.19, 20.14) |
|  | June | 1000 | | 11.10 (9.22, 13.21) | 10.80 (8.94, 12.89) | | 859 | 15.24 (11.33, 20.19) | 14.97 (11.08, 19.93) |
|  | July | 999 | | 10.51 (8.68, 12.58) | 10.31 (8.49, 12.36) | | 874 | 14.59 (10.97, 19.14) | 14.45 (10.87, 18.96) |
|  | August | 3664 | | 10.02 (9.06, 11.03) | 9.74 (8.80, 10.75) | | 3087 | 15.88 (13.83, 18.16) | 15.73 (13.68, 18.03) |
| **San Francisco** |  |  | |  |  | |  |  |  |
|  | March | 999 | | 0.10 (0.00, 0.56) | 0.10 (0.00, 0.56) | | 718 | 0.11 (0.01, 0.82) | 0.11 (0.01, 0.82) |
|  | April | 1000 | | 0.80 (0.35, 1.57) | 0.30 (0.06, 0.87) | | 720 | 1.04 (0.35, 3.03) | 0.15 (0.03, 0.76) |
|  | May | 988 | | 1.01 (0.49, 1.85) | 0.81 (0.35, 1.59) | | 729 | 0.57 (0.28, 1.17) | 0.44 (0.19, 1.01) |
|  | June | 999 | | 0.60 (0.22, 1.30) | 0.50 (0.16, 1.16) | | 716 | 1.13 (0.36, 3.49) | 1.13 (0.36, 3.49) |
|  | July | 2000 | | 1.10 (0.69, 1.66) | 1.00 (0.61, 1.54) | | 1421 | 0.98 (0.57, 1.68) | 0.91 (0.51, 1.62) |
|  | August | 2000 | | 1.55 (1.06, 2.19) | 1.45 (0.97, 2.08) | | 1404 | 1.48 (0.70, 3.11) | 1.48 (0.70, 3.11) |
| **Seattle** |  |  | |  |  | |  |  |  |
|  | March | 1000 | | 0.30 (0.06, 0.87) | 0.20 (0.02, 0.72) | | 922 | 0.25 (0.08, 0.80) | 0.18 (0.04, 0.77) |
|  | April | 1000 | | 0.90 (0.41, 1.70) | 0.70 (0.28, 1.44) | | 819 | 1.23 (0.54, 2.74) | 1.01 (0.39, 2.60) |
|  | May | 1003 | | 1.00 (0.48, 1.83) | 1.00 (0.48, 1.83) | | 835 | 0.97 (0.43, 2.16) | 0.97 (0.43, 2.16) |
|  | June | 1000 | | 1.60 (0.92, 2.59) | 1.30 (0.69, 2.21) | | 836 | 2.00 (0.89, 4.44) | 1.75 (0.69, 4.34) |
|  | July | 2008 | | 1.74 (1.22, 2.42) | 1.44 (0.97, 2.07) | | 1704 | 1.60 (1.08, 2.36) | 1.40 (0.90, 2.17) |
|  | August | 2008 | | 2.29 (1.68, 3.04) | 2.09 (1.51, 2.82) | | 1682 | 2.28 (1.52, 3.42) | 1.87 (1.22, 2.87) |
| **Boston** |  |  | |  |  | |  |  |  |
|  | April | 999 | | 0.80 (0.35, 1.57) | 0.50 (0.16, 1.16) | | 944 | 1.12 (0.41, 3.03) | 0.86 (0.24, 3.06) |
|  | May | 1000 | | 1.40 (0.77, 2.34) | 1.30 (0.69, 2.21) | | 859 | 1.37 (0.70, 2.67) | 1.37 (0.70, 2.67) |
|  | June | 1000 | | 1.80 (1.07, 2.83) | 1.60 (0.92, 2.59) | | 941 | 2.50 (1.35, 4.56) | 2.30 (1.19, 4.41) |
|  | July | 2000 | | 2.90 (2.21, 3.73) | 2.60 (1.95, 3.40) | | 1744 | 3.99 (2.91, 5.46) | 3.78 (2.71, 5.25) |
|  | August | 2000 | | 3.55 (2.78, 4.46) | 3.20 (2.47, 4.07) | | 1727 | 4.56 (3.21, 6.45) | 4.21 (2.93, 6.02) |
| **Los Angeles** |  |  | |  |  | |  |  |  |
|  | April | 1000 | | 0.90 (0.41, 1.70) | 0.80 (0.35, 1.57) | | 882 | 0.91 (0.39, 2.11) | 0.79 (0.30, 2.04) |
|  | May | 1000 | | 0.60 (0.22, 1.30) | 0.50 (0.16, 1.16) | | 863 | 0.68 (0.25, 1.82) | 0.68 (0.25, 1.82) |
|  | June | 1000 | | 1.20 (0.62, 2.09) | 1.10 (0.55, 1.96) | | 857 | 1.65 (0.82, 3.32) | 1.61 (0.78, 3.30) |
|  | July | 4000 | | 2.15 (1.72, 2.65) | 1.98 (1.57, 2.46) | | 3215 | 2.50 (1.82, 3.42) | 2.39 (1.71, 3.32) |
|  | August | 4000 | | 3.38 (2.84, 3.98) | 3.25 (2.72, 3.85) | | 3303 | 4.53 (3.84, 5.34) | 4.50 (3.81, 5.30) |
| **Minneapolis** |  |  | |  |  | |  |  |  |
|  | April | 1000 | | 0.40 (0.11, 1.02) | 0.20 (0.02, 0.72) | | 830 | 0.78 (0.17, 3.42) | 0.67 (0.12, 3.74) |
|  | May | 1000 | | 0.80 (0.35, 1.57) | 0.50 (0.16, 1.16) | | 861 | 0.76 (0.30, 1.92) | 0.54 (0.16, 1.76) |
|  | June | 1000 | | 0.80 (0.35, 1.57) | 0.60 (0.22, 1.30) | | 868 | 0.81 (0.30, 2.21) | 0.69 (0.22, 2.17) |
|  | July | 2000 | | 1.25 (0.81, 1.84) | 1.05 (0.65, 1.60) | | 1745 | 1.87 (0.90, 3.83) | 1.78 (0.83, 3.79) |
|  | August | 2000 | | 1.60 (1.10, 2.25) | 1.25 (0.81, 1.84) | | 1747 | 1.80 (1.17, 2.74) | 1.47 (0.93, 2.33) |

**Supplemental Table 2**. Comparison of weighted seroprevalence (point estimate and 95% CI) determined by Ortho CoV2T reactivity versus weighted seroprevalence determined by reactive results in the supplementary testing algorithm by region and month. 95% CI = 95% confidence interval; DCR = donor catchment region

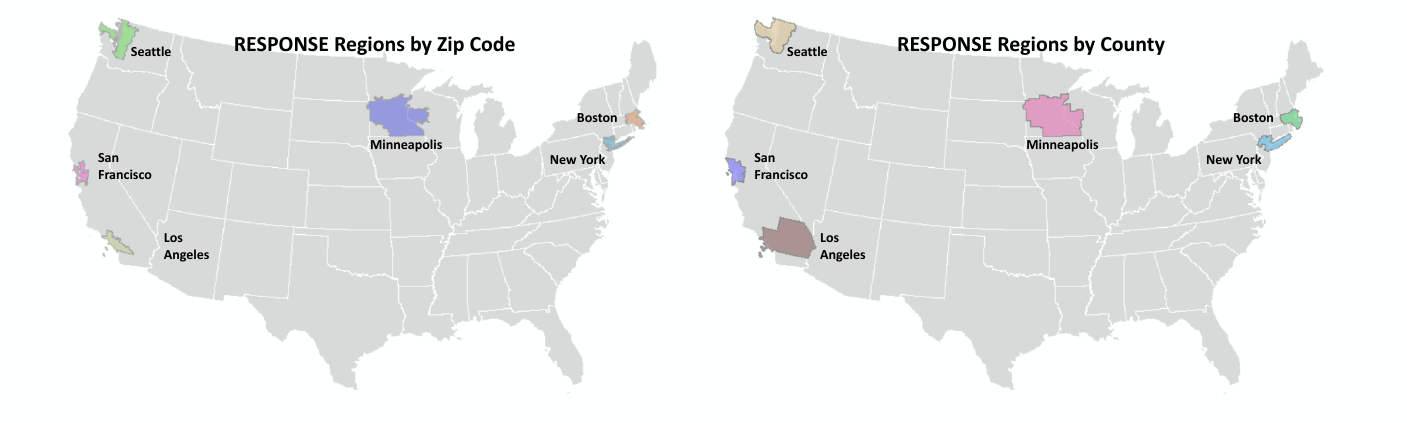

**Supplemental Figure 1:** Map of DCRs- showing regions -ZIP3 and by county. DCR = donor catchment region

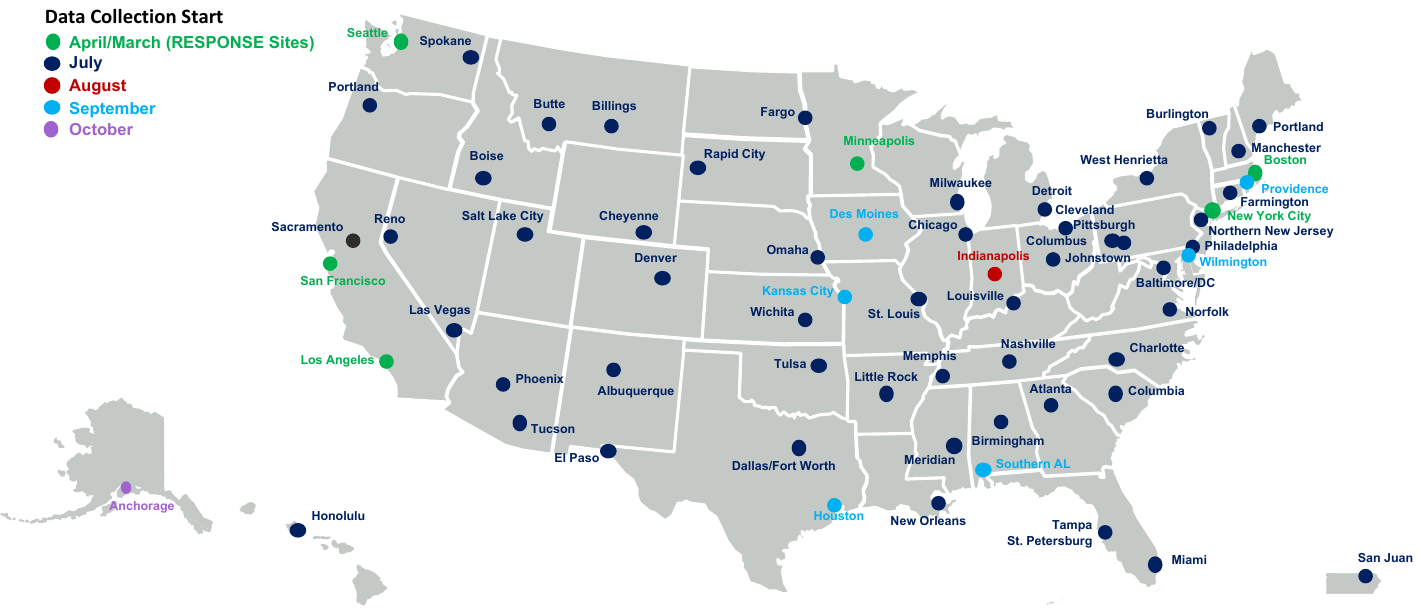

**Supplemental Figure 2:** Map of the RESPONSE regions and the additional 56 regions that are included in the national CDC blood Donor serosurvey. Color code indicates the month that data accrual began.

**Appendix A. Laboratory Methods**

*Ortho VITROS S1 Total Ig Assay for initial screening*

Serum samples were tested to measure anti-SARS-Cov2 total Ig level on the Ortho Vitros 3600 (Vitros CoV2T) at Vitalant Research Institute San Francisco (VRI-SF) and Creative Testing Solutions (CTS), following the manufacturer’s instructions. This test allows identification of total antibodies (IgG, IgM, IgA) against the SARS-CoV-2 virus S1 spike protein and was released under EUA (the first SARS-CoV-2 serology assay to receive EUA).(1) Briefly, in this chemiluminescence assay, antibodies present in the serum are bound in a double sandwich configuration to the SARS-CoV-2 S1 spike protein antigen on the testing wells and to horseradish peroxidase (HRP)-labeled recombinant SARS-CoV-2 antigen in the liquid phase. HRP catalyzes the light signal that is captured by the instrument’s luminometer and classified as negative or positive, based on a threshold Signal/Cutoff (S/C) of 1.0. Based on the FDA EUA IFU the assay has 100% specificity based on testing normal donor samples and 100% sensitivity for samples collected >8 days after COVID-19 symptom onset. Based on distribution of S/C values the assay has a wide dynamic range with reactive results ranging from 1–1000, and testing can be performed with high throughput on multiple Vitros platforms.

*Roche NC Total Ig Assay*

The Elecsys Anti-SARS-CoV-2 (Elecsys CoV2T) assay is a qualitative, serological, double-antigen sandwich principal immunoassay to be used on the **cobas e** immunoassay family of analyzers with an 18-minute test time. The Elecsys Anti-SARS-CoV-2 assay uses a recombinant protein representing the nucleocapsid (N) antigen (i.e. full length nucleocapsid protein) for the determination of antibodies against SARS-CoV-2. Both labeled antigens have the same antigen amino acid sequence. The antigen is presented in a way to capture predominantly anti-SARS-CoV-2 IgG, but also anti-SARS-CoV-2 IgA and mature anti-SARS-CoV-2 IgM.

Anti-SARS-CoV-2 specific antibodies are captured to the streptavidin coated solid phase microparticles with biotinylated SARS-CoV-2 specific antigen, the ruthenylated antigen mediates detection of the complex via electrochemiluminescence. The double-antigen sandwich complex is magnetically captured onto an electrode and the bound complex is washed. Application of voltage to the electrode induces chemiluminescence, which is measured by a photomultiplier tube. Results are determined via a 2-point calibration and a cutoff formula encoded in the reagent barcode. The testing principle is shown in Figure 1 below.

**Testing Principle of Elecsys Anti-SARS-CoV-2**

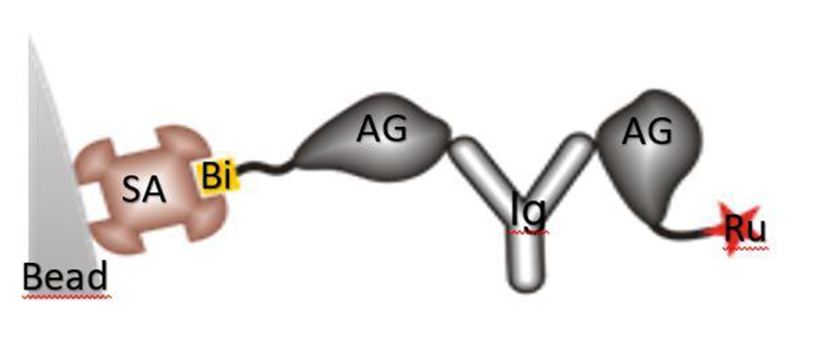

1st incubation: 20 μL of sample (**cobas e** 411, **cobas e** 601, and **cobas e** 602 analyzers) or 12 μL of sample (**cobas e** 801 analyzer), biotinylated SARS-CoV-2 specific recombinant antigen and SARS-CoV-2 specific recombinant antigen labeled with a ruthenium complex^a)^ react to form a sandwich complex.
^a)^ Tris(2,2'-bipyridyl)ruthenium(II)-complex (Ru(bpy))

2nd incubation: After addition of streptavidin-coated microparticles, the complex becomes bound to the solid phase via interaction of biotin and streptavidin. The reaction mixture is aspirated into the measuring cell where the microparticles are magnetically captured onto the surface of the electrode. Unbound substances are then removed with ProCell/ProCell M/ProCell II M. Application of a voltage to the electrode then induces chemiluminescent emission, which is measured by a photomultiplier. Results are determined automatically by the software by comparing the electrochemiluminescence signal obtained from the reaction product of the sample with the signal of the cutoff value previously obtained by calibration. The sensitivity and specificity are 65.5–100% and 99.81%, respectively (2). The results are reported qualitatively as positive or negative for SARS-COV2.

*Pseudovirus-based Reporter Virus Neutralization (RVPN)*

CoV-2 seroreactive samples were tested with the SARS-CoV-2 Reporter Viral Particle Neutralization (RVPN) assay (PMID: 32943630) at VRI. Upon receipt serum samples were plated in a 96-well plate and heat-inactivated (HI) for 30 min at 56°C. Reporter viral particles (RVPs) represent a safe and rapid way of quantitatively measuring neutralization, by using SARS-CoV-2 Spike glycoprotein pseudotyped onto a rhabdovirus reporter virus. Neutralization was performed using vesicular stomatitis virus (VSV)-based pseudovirions as previously described. (3) Briefly, HEK293T target cells stably expressing human ACE2 and TMPRSS2 were plated into 96-well plates. Heat inactivated serum was mixed with VSV/SARS-CoV-2 Spike pseudovirus harboring firefly luciferase reporter and incubated for 1 hour at 37oC and then used to infect the target cells. After 18-24 hours at 37°C, supernatant was removed, cells lysed and luciferase activity was measured as per manufacturer’s instructions (Promega, Madison). Greater than 50% reduction in signal compared to a no serum control was considered positive.

**Appendix B. Statistical Methods**

**B1. Data Collection (Selection of monthly samples for antibody testing)**

The samples of blood donations were selected monthly by the blood collection centers. The monthly sample sizes were typically 1000 donations and for selected centers increased to 2000 or 4000 in July or August (see Appendix Table B2). No formal randomization procedures were used, with the center laboratories given latitude in determining samples to be tested (the laboratory sees the donation aliquot and knows nothing about demographics of the donor). This latitude was deemed necessary, as the laboratory’s main function is to provide products for transfusion, and any methodology to ensure a more random sample may have adversely affected this function. Typically, the samples were composed of consecutive donations received by each center’s laboratory early in the month. COVID-19 convalescent plasma (CCP) donations were excluded. Monthly center sampled donation demographics were compared to monthly center total donation demographics to assess representativeness of monthly sampled donations to monthly total donations via chi-square statistics.

**B1.1. Representativeness of samples selected and tested for SARS-CoV-2 antibodies relative to all donations**

In total, 499,476 non-COVID convalescent plasma donations were collected in all participating regions during the study period, of which 50,156 (10%) were included in the study. A comparison of the demographic characteristics of all donors to those sampled did not show any consistent month-to-month differences. Although differences were found in the monthly regional percentages of first-time versus repeat donors, these varied in direction and might have occurred because of small monthly sample sizes. (Table 1, supplemental Table S1).

B2. Donation Catchment Region (DCR)

Because the goal of the study is to estimate the seroprevalence among the general population, not among the donor population, two adjustments were applied to the monthly sample data. The first is the determination of a geographic area that is well represented by the monthly samples and the second is a weighting adjustment to account for demographic differences between the donors and the general population. The first adjustment is described here, while the second is explained in Appendix section B3.

For each of the six regions, a “donation catchment region” (DCR) was derived based on the ZIP code frequencies of the total center donations in March, using an iterative process. An initial DCR is created from the center’s own 3-digit zip code and all neighboring 3-digit zip codes. Then, the most common next 3-digit zip code was added to the DCR (plus any other 3-digit zip codes to ensure the DCR remains a contiguous region). This is repeated until at least 80% of the March total donations were included. Some minor exceptions were allowed; for instance the Boston region was constrained to be wholly within Massachusetts. And the San Francisco region did not attain the 80% goal, since to achieve that goal, Santa Rosa donors would have been included (Santa Rosa was deemed too distant to be logically interpreted as part of the San Francisco region).

The process was repeated using April total donations to determine the stability of the DCR over time, after which they were kept fixed for the remainder of the study. Appendix Table B2 shows the monthly percentages of DCR samples among samples.

Appendix Table B2. Donations, Samples, and DCR Samples within region by Center and month.

| Center | Month | Donations | Samples | DCR Samples | DCR samples/ samples (%) |
| --- | --- | --- | --- | --- | --- |
| New York | March | 28432 | 1493 | 1331 | 89% |
|  | April | 15505 | 991 | 874 | 88% |
|  | May | 20114 | 985 | 854 | 87% |
|  | June | 25565 | 1000 | 859 | 86% |
|  | July | 21003 | 999 | 874 | 87% |
|  | August | 22103 | 3664 | 3087 | 84% |
| San Francisco | March | 4342 | 999 | 718 | 72% |
|  | April | 3840 | 1000 | 720 | 72% |
|  | May | 3792 | 988 | 729 | 74% |
|  | June | 6144 | 999 | 716 | 72% |
|  | July | 5670 | 2000 | 1421 | 71% |
|  | August | 4970 | 2000 | 1404 | 70% |
| Seattle | March | 13972 | 1000 | 922 | 92% |
|  | April | 9831 | 1000 | 819 | 82% |
|  | May | 13304 | 1003 | 835 | 83% |
|  | June | 12053 | 1000 | 836 | 84% |
|  | July | 14041 | 2008 | 1704 | 85% |
|  | August | 13008 | 2008 | 1682 | 84% |
| Boston | March | 10400 | n/a | n/a | n/a |
|  | April | 7344 | 999 | 944 | 94% |
|  | May | 8336 | 1000 | 859 | 86% |
|  | June | 10327 | 1000 | 941 | 94% |
|  | July | 10939 | 2000 | 1744 | 87% |
|  | August | 10491 | 2000 | 1727 | 86% |
| Los Angeles | March | 24661 | n/a | n/a | n/a |
|  | April | 17242 | 1000 | 882 | 88% |
|  | May | 21435 | 1000 | 863 | 86% |
|  | June | 25647 | 1000 | 857 | 86% |
|  | July | 26738 | 4000 | 3215 | 80% |
|  | August | 23630 | 4000 | 3303 | 83% |
| Minneapolis | March | 21083 | n/a | n/a | n/a |
|  | April | 15400 | 1000 | 830 | 83% |
|  | May | 16873 | 1000 | 861 | 86% |
|  | June | 21538 | 1000 | 868 | 87% |
|  | July | 22820 | 2000 | 1745 | 87% |
|  | August | 21379 | 2000 | 1747 | 87% |

Table 4 compares DCR weighted seroprevalences with cases as reported to CDC. However, cases reported to CDC are by county of residence, and zip code is unknown. Hence, in order to make this comparison meaningful, it is necessary to define a county-based approximation to the DCR, referred to as the “county-based DCR” in what follows. For each DCR, the corresponding county-based DCR was created using 2018 American Community Survey (ACS) data with the following rule: counties for which the sum of DCR’s overlapping ZIP codes comprised ≥30% of the county population were included in the county-based DCR, while those with less than 30% were excluded. Supplemental Figure 3 shows the correspondence between the ZIP code and county based DCRs.

B3. Weighting

Following creation of the DCRs, a weighting adjustment was used to account for demographic differences between the sample donors and the general population. The adjustment consists of creating “estimation weights” for the individual sample donations, so that their over- or underrepresentation in different demographic groups relative to the general population is adjusted. This approach is sometimes referred to as a “pseudo-design” and is the most commonly used statistical method to estimate general population characteristics based on nonprobability data (Elliott and Valliant, 2017).

The monthly estimation weights were obtained by raking using the 2018 American Community Survey (ACS) estimates for the age, gender, and race/ethnicity composition for the DCRs. Raking (Oh and Scheuren, 1987) is a statistical calibration technique frequently used in large-scale surveys. In addition to these estimation weights, monthly sets of 50 replicate weights were created (Rust and Rao, 1996), which were used to estimate the variance of the weighted seroprevalence estimates.

B4. Analysis

When using these monthly estimation weights and sample data in the DCR, the resulting weighted seroprevalence estimates can be interpreted as statistically valid estimates of the general population monthly seroprevalence in the DCR, with their standard error and confidence interval computable using the replicate weights. Weighted seroprevalence estimates for specific demographic groups can likewise be computed by subsetting the data to the relevant groups. Figure 2 shows each center’s seroprevalence over time, and Table 2 shows each center’s seroprevalence in August by demographics.

An estimated total number of cumulative infections for a county-based DCR was computed as the ZIP-code–based DCR seroprevalence multiplied by the Census data county-based DCR total (county-based DCR and ZIP-code–based DCR seroprevalences are assumed equal; a seemingly reasonable assumption since the corresponding DCR pairs greatly overlap as shown in Supplemental Figure 3). Table 4 and Figure 3 presents seroprevalence, estimated cumulative infections, cases reported (by county-based DCRs).

Since seroprevalence in the U.S. population is known to vary by location and time, a stratified (by center and month) logistic regression model was developed to assess demographic effects. The model results are shown in Table 3. More complex models were investigated (e.g. a temporal age effect was postulated; i.e. young donors may have lower seroprevalence early in the pandemic and higher seroprevalence later). However, the total number of tested donations was not large enough to find any statistically significant interactions.

**Appendix C. Development and validation of supplemental testing algorithm**

Results for 483 of 489 Vitros CoV2T reactive donation samples identified from screening 50,156 donations from March–June 2020 that were tested in parallel by the Elecsys CoV2T and RVNPT assays are shown in **Figure 1**(six samples had insufficient volume for Elecsys CoV2T and RVPNT testing). We stratified Vitros CoV2T reactivity by signal to cutoff (S/C) ratios of 1–10 vs ≥10. Of 79 samples with Vitros CoV2T S/C 1.0 to <10, 22 (28%) were confirmed by both the Elecsys CoV2T and the RVNPT assays, 12 (15%) were reactive on the Elecsys CoV2T but negative for the RVPNT assay, seven (9%) were Elecsys CoV2T non-reactive but had positive NT50 titers (ranging from 40 to 640) and 38 (48%) were nonreactive by both supplemental assays. In contrast, 367 (91%) of 404 samples with Vitros CoV2T S/C≥10 tested reactive by both the Elecsys CoV2T and RVPNT assays, while 19 samples (4.7%) were only reactive on the Elecsys CoV2T assay and 17 samples (4.2%) were only reactive by RVPNT. Only one sample (0.02%) with Vitros CoV2T S/C ≥10 tested negative by both the Elecsys CoV2T and RVPNT assays. Thus, we modified the supplemental testing algorithm from July onward to restrict supplemental testing to samples with Vitros CoV2T S/C 1–10, with those samples first tested by the Elecsys CoV2T followed by reflex testing of non-reactive samples by RVPNT. Samples were classified as “confirmed antibody positive” if: i) they had a S/C ≥10 on Vitros CoV2T screening assay, thus no futher testing was needed; or ii) if the S/C was 1–10 and they tested reactive on either the Elecsys CoV2T or RVNPT assay.(Figure 1, Panel B)

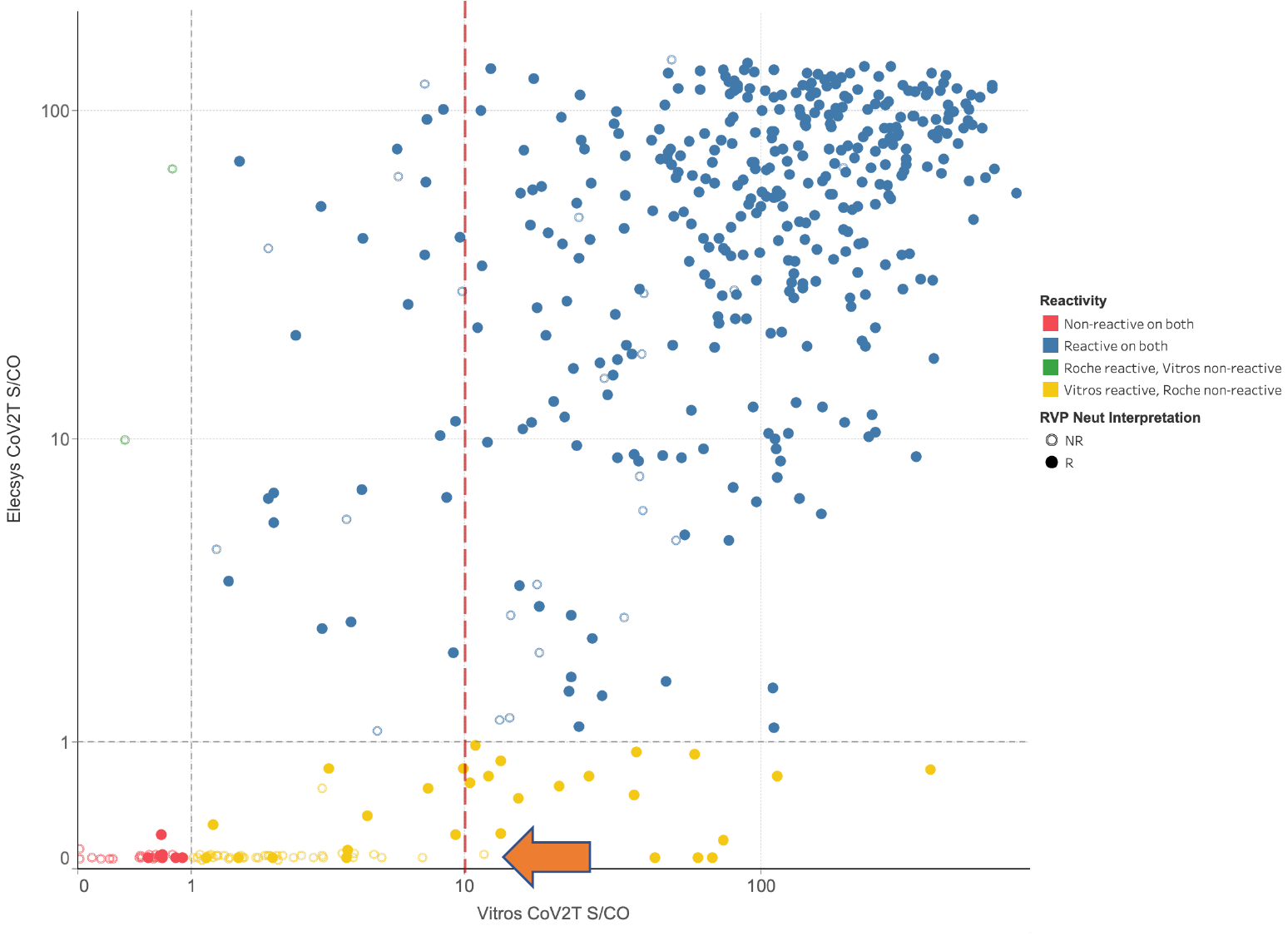

**Supplemental Figure 5.** Scatter plot of S/CO ratios of Vitros S1 Total Ig CoV2T assay (x axis) and Roche NC Total Ig assay (y axis). Confirmatory testing status was determined based either on the Roche assay results or the pseudovirus neutralization results (RVPN). Blue: Vitros CoV2T, Elecsys CoV2T reactive; Red: Vitros CoV2T, Elecsys CoV2T non reactive; green: Vitros CoV2T non-reactive, Elecsys CoV2T reactive; yellow: Vitros CoV2T reactive, Elecsys CoV2T non-reactive. Open circles; RVPN nonreactive; closed circles RVPN reactive. Orange arrow indicates on sample with Vitros CoV2T S/C ≥10 tested negative by both the Elecsys CoV2T and RVPNT assays.
